# Machine Learning Classification Of Autism Spectrum Disorder Based On Reciprocity In Naturalistic Social Interactions

**DOI:** 10.1101/2022.12.20.22283571

**Authors:** J.C. Koehler, M.S. Dong, A.M. Nelson, S. Fischer, J. Späth, I.S. Plank, N. Koutsouleris, C.M. Falter-Wagner

## Abstract

Autism spectrum disorder is characterized by impaired social communication and interaction. As a neurodevelopmental disorder typically diagnosed during childhood, diagnosis in adulthood is preceded by a resource-heavy clinical assessment period. The ongoing developments in digital phenotyping give rise to novel opportunities within the screening and diagnostic process. In this study, we investigated videos of naturalistic social interaction between autistic and non-autistic adults on their predictiveness for autistic behaviors. Non-autistic control participants were either paired with each other or an autistic participant to engage in two conversational tasks. We used existing computer vision algorithms to extract information based on the synchrony of movement and facial expression. These were subsequently used as features in a support vector machine learning model to predict interaction dyad membership. Results showed high predictive accuracy of synchrony in facial movements, underlining the distinctive nature of non-verbal behavior in autism and its feasibility for digitalized assessment.

## 1. Introduction

The diagnosis of autism spectrum disorder (ASD) encompasses a range of symptoms in reciprocal social interaction and communication as well as restricted, repetitive behaviors and interests ^1^. The currently rising prevalence ^2^ exacerbates waiting times for an already long and demanding diagnostic process, increasing psychological stress on seeking diagnostic clarification ^3^. Gold-standard recommendations include assessment with semi-structured diagnostic interviews or observational tools conducted by multidisciplinary teams, along with neuropsychological assessments and an anamnesis of developmental history by a caregiver ^3^. With the increasing number of patients seeking diagnosis in adulthood, the lack of recommended diagnostic instruments for this population ^4^ poses an additional challenge.

Digitalized methods have high potential to improve screening and diagnostic procedures, such as assessing home videos ^5^ or interactions with virtual characters ^6,7^. While promising, these findings often rely on time-consuming manual behavioral coding or, more importantly, may not adequately reflect real-time social interactions, which are especially relevant for judging symptom strength ^8^. Additionally, the increased use of artificial intelligence methods, such as machine learning (ML), has furthered research on increasing the efficiency of existing diagnostic tools, e.g., by identifying subsets of the most important items for diagnosis ^9,10^. While these results are prone to a certain circularity, introduced by the dependency of features used for classification and composition of diagnostic groups in the first place, they nevertheless point to areas of impairments most indicative for diagnosis. These include aberrances in, e.g., gesturing, facial expressions and reciprocal social communication ^9^. These traits seem to influence first impressions of people with ASD, who are judged as interacting more awkwardly by typically developing (TD) peers ^11^. Importantly, this even holds true for the judgements of short, non-verbal excerpts of behavior ^12^, suggesting that non-verbal behavior represents an important pillar of clinical impression formation.

A way to quantify this aberrant interaction style is through closely examining the way two interacting partners temporally adjust their behavior with each other, or, in other words, how well they are “in sync”. Interpersonal synchrony or coordination can not only be achieved through mutual, bilateral matching, but also by establishing leader-follower relationships through unilaterally adapting to the behavior of the interactant ^13^. Interpersonal synchrony has repeatedly been associated with rapport, affiliation, and perception ^14,15^, emphasizing its importance for social cognition. In ASD, reduced interpersonal synchrony or coordination has in fact been described on multiple modalities and across the lifespan ^16^. For instance, autistic adults exhibit less mimicry in response to others’ dynamic facial expressions ^17^. Reduced coordination of emotional facial expressions has also been found in autistic youth in conversation with a partner ^18^. This affect coordination has also been investigated in the context of diagnostic classification, showing high accuracy in a standardized social interaction task with a virtual character ^6^. Interpersonal synchrony in head motion has been found to be reduced in diagnostic interviews with patients subsequently diagnosed with autism as compared to those who were not ^19^. Another study investigating head and body motion synchrony in autistic and non-autistic adults found both to be reduced when an autistic person was part of the conversation ^20^, once again reflecting the importance of the interactional perspective. Further, synchrony and coordination differences in autism have also been found within the individual (intrapersonally), with reduced coordination of simultaneous movements ^21,22^ or different communication modalities ^23^. A pronounced example are motor stereotypies, such as repeated movements of isolated body parts (e.g. hand flapping or tapping ^24^). In a study assessing the coordination between head and body within conversations between autistic and non-autistic individuals, a Support Vector Machine (SVM) model showed classification of autism with an accuracy of 75.9% ^25^. Lastly, movement atypicalities, apart from coordination, appear to be pronounced in autism, both for facial expressiveness and full body movement. For example, autistic children were found to be less expressive than TD children when rated by adults naïve to the diagnosis ^26^. Additionally, biological motion and motor control has been found to compose a unique kinematic profile for autism ^27^. A recent meta-analysis found a significant correlation between gross motor and social skills in autism ^28^, underlining the significance of movement differences for the core symptomatic profile of ASD.

In summary, the mere definition of ASD as a disorder of social interaction implies an interdependency and calls for shifting to the interactional dyad as unit of analysis ^29^. However, feasible measures are lacking due to their reliance on extensive manual coding, experimental paradigms appearing staged or unnaturalistic, or investigating only isolated aspects of social interaction.

Thus, the aim of this proof-of-concept study was to build upon existing knowledge of adaptation difficulties in autism and use the richness of non-verbal social interaction data in an efficient way to build an objective (i.e. independent of self-or clinician-ratings) classification model of autistic social interaction. To this end, we trained several machine learning classification models to optimally differentiate between members of autistic vs. non-autistic interactional dyads. To increase objectivity and feasibility for potential further development in clinical practice, we used existing open-source algorithms that maximized automation in the annotation and analysis process.

## 2. Results

Results are described in four sections. First, we describe results of our base classification models based on the respective adaptation of facial expression, head movement, and body movement of the participants to their partner, as well as the intrapersonal head-body coordination, and global movement parameters. Participants were classified as belonging to a mixed (ASD-TD) vs. non-autistic control (TD-TD) dyad. We then present clinical characteristics of our autistic participants in our top-performing model and in which way they differed depending on the group they were classified into. Subsequently, the results for our combined classification models are described. Finally, we summarize our findings on classification based on diagnostic group and compare it to our dyad-based classification approach. A summary of the most important SVM metrics for all models is depicted in Figure 4. Further details on feature extraction and analysis procedures can be found in the supplementary information.

### 2.1. Base Model Performances

Using facial action unit (AU) synchrony data, the repeated nested stratified cross-validation FACEsync model yielded a balanced accuracy (BAC) of 79.5%, and an area under the curve (AUC) of .82 (model significance *p* < .001). The contribution of the different features to classification group was calculated by feature weights (see Supplementary Information S3.4) and cross-validation ratio (Figure 1). Additionally, the sign-based consistency was explored as an indicator of the feature classification reliability (Figure 2). Assignment to the ASD-TD dyads was mainly driven by features describing an elevated or highly varied extent of adaptation in AU17 (chin raiser) and AU26 (jaw drop). Minimized adaptation in AU01 (inner brow raiser), AU20 (lip stretcher) and AU45 (blink) were indicative of belonging to the TD-TD interaction type. In order to investigate any associations of facial emotion recognition abilities and adaptation behaviors of the different facial AUs, correlation analyses were performed between the decision scores derived from the FACEsync model and accuracy and response time (rt) from the Berlin Emotion Recognition Test (BERT ^30^). No significant associations were found (*r*_*accuracy*_(86) = -.16, *r*_*rt*_(86) = .13; both *p* = .23 after FDR correction).

**Figure 1.**
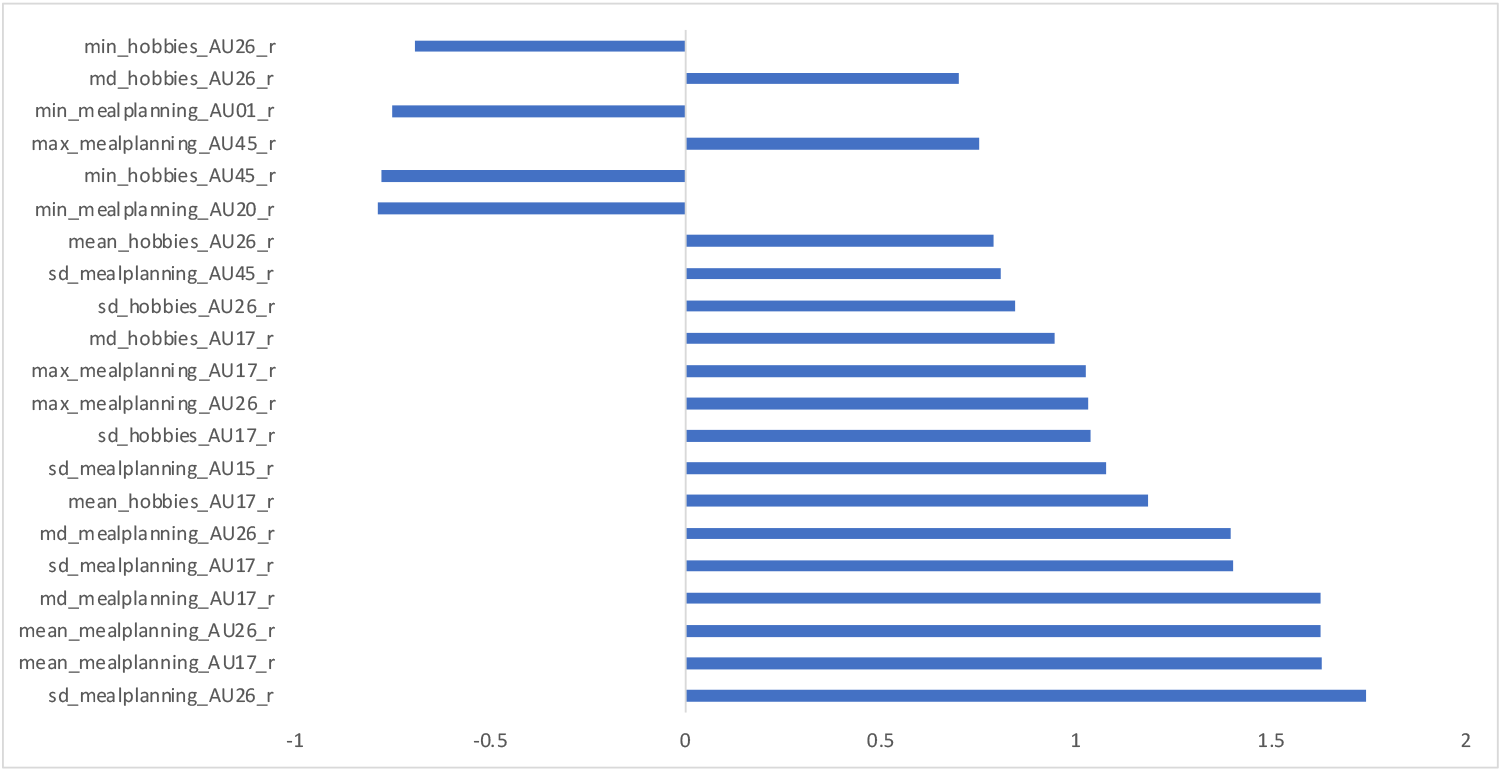
Cross-validation ratio of feature weights for FACEsync model.

**Figure 2.**
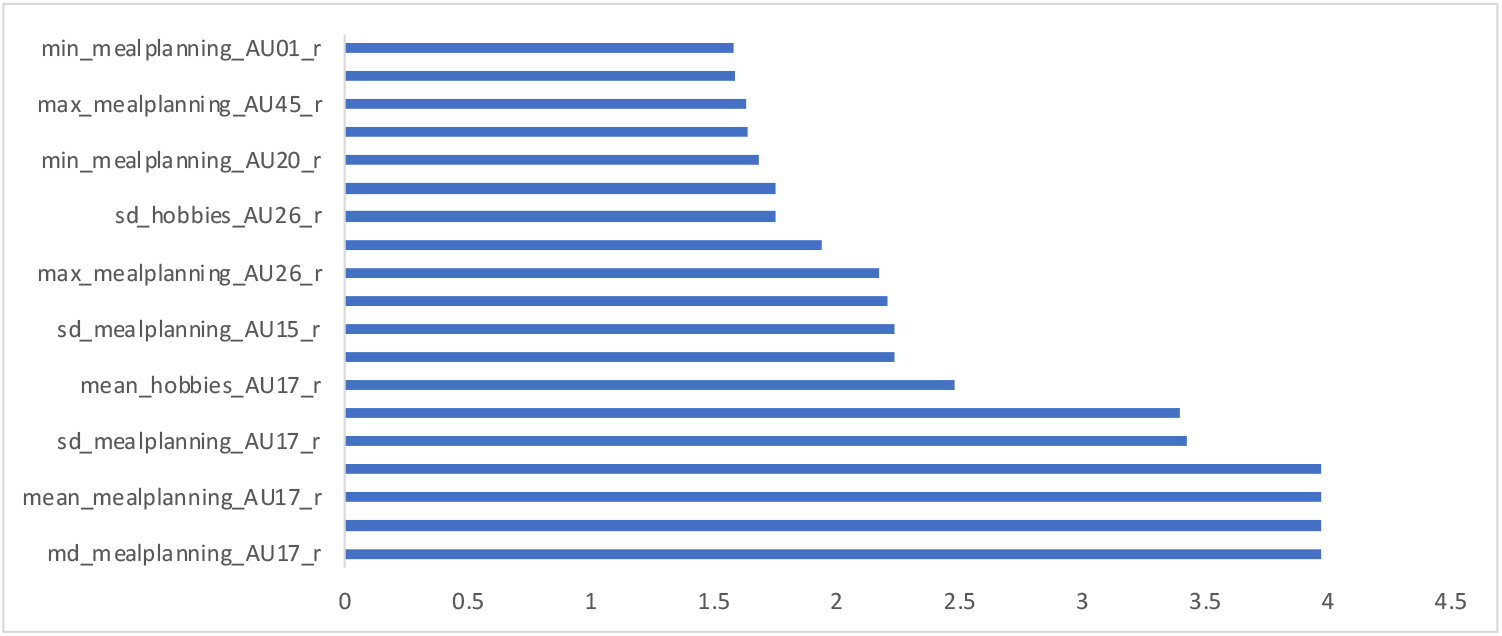
Sign-based consistency (log-transformed, p-value FDR-corrected) for FACEsync model.

The model using only head motion coordination data (HEADsync) achieved a BAC of 62.1% and an AUC of .64 (model significance *p* = .002). Assignment to the TD-TD group was driven by higher values in minimum adaptation of global head movement whereas higher maximum and more variant values for head movement adaptation predicted the ASD-TD group.

The classification model based on upper body movement coordination (BODYsync) predicted dyad origin around chance level with a BAC of 56.7% and an AUC of .55 (model significance *p* = .011).

Our classification model based on intrapersonal head-body coordination (INTRAsync) performed around chance level with a BAC of 44.2% and an AUC of .44 (model significance *p* = .999).

The SVM classification model based on features of total head and body movement and general facial expressiveness (MovEx) predicted dyad origin with a BAC of 68.8% and an AUC of .75 (model significance *p* < .001).

Additional classification metrics for all models can be found in Supplementary Table S5.

### 2.2. Clinical characteristics of correctly vs. incorrectly classified autistic participants

We were interested whether there were any notable differences in clinical characteristics of the autistic participants within our ASD-TD interactions who were classified correctly (true positive, TP) vs. incorrectly (false negative, FN) as belonging to a non-autistic control dyad. For this, we ran a series of Welch independent sample two-sided t-tests within the autistic subsample for all base classification models. Figure 3 depicts the group differences for the FACEsync model. While TP autistic participants on average had lower alexithymia scores (*M* = 57.91) than FN autistic participants (*M* = 86.60), this difference did not survive FDR correction (*p* = .07). We found no significant differences between autistic participants for the other models (see Supplementary Tables S6-10).

**Figure 3.**
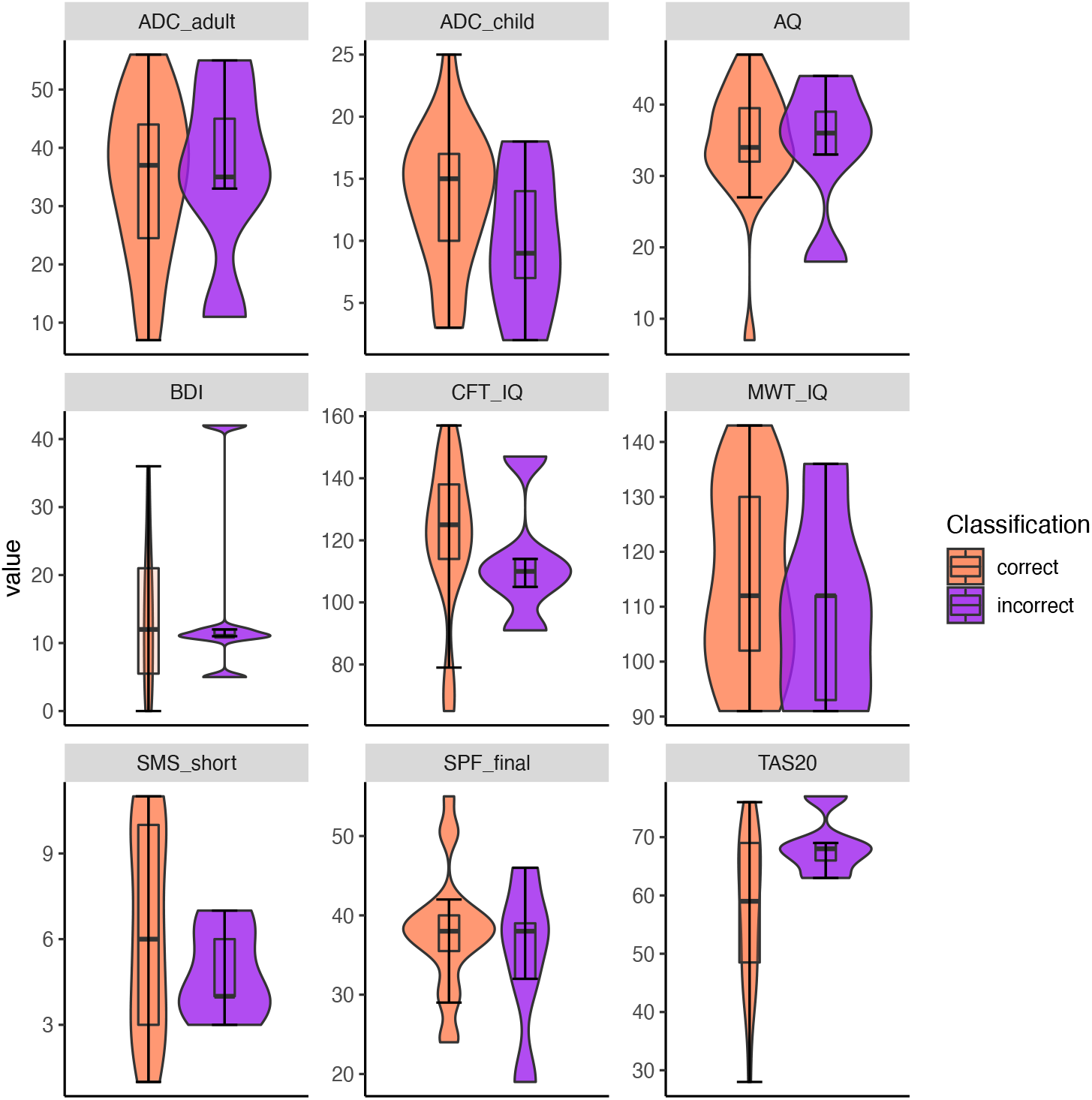
Clinical characteristics of TP vs. FN autistic individuals in FACEsync model.

**Figure 4.**
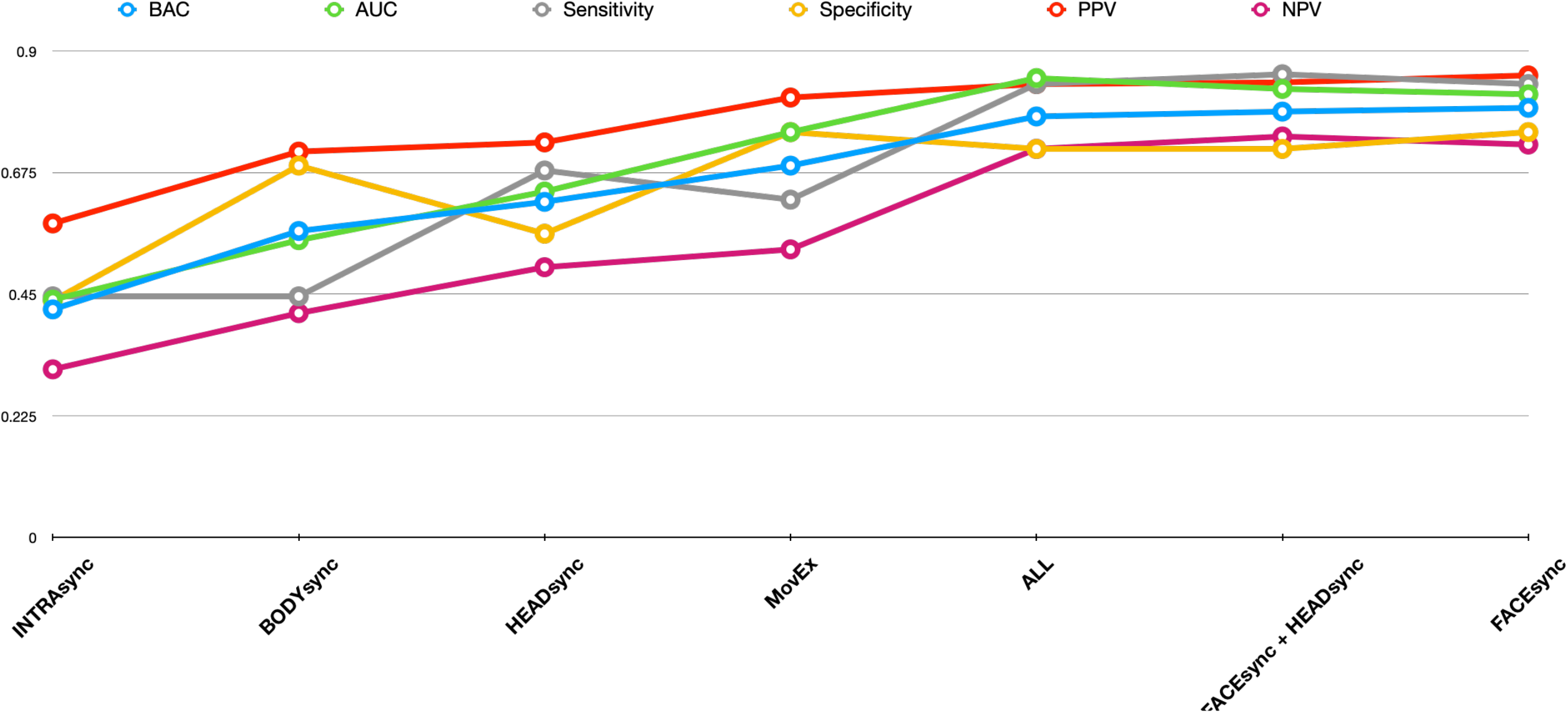
Classification metrics for all base and stacking models.

### 2.3. Stacking Model

All base model decision scores were extracted and combined into a hierarchical stacking-based fusion framework to assess potential prediction improvements. Combinations of only the head region (FACEsync + HEADsync; BAC = 78.8%, AUC = .83), as well as of all modalities (BAC = 77.9%, AUC = .85) did not outperform the most predictive base model (FACEsync) with 79.5%.

### 2.4. Classification based on diagnostic group

We repeated all SVM analyses using different labels based on diagnostic groups while ignoring interaction type. These additional analyses were conducted in order to investigate if our collected social interaction data was specific enough to identify an autistic individual, regardless of interaction dyad origin. All models generated inferior prediction accuracies compared to the dyad labelling approach (2.1). Detailed results can be found in Supplementary Information S3.6.

## 3. Discussion

The aim of the current study was to quantify social interaction in ASD for the purpose of automatized diagnostic classification. In this proof-of-concept study, we set out to utilize a dyadic setting for classification of autistic vs. non-autistic interaction based on reciprocity. Participants were filmed conducting two brief conversations about pre-set topics. Using repeated nested cross-validation techniques, we could show that SVM classification models based on different modalities of behavioral reciprocity were sufficient to predict dyad membership to a high degree. Contrary to our hypothesis, combining different non-verbal modalities did not improve overall predictive accuracy. Classification into individual diagnostic groups (ASD vs. TD) based on social interaction data performed worse on all modalities, as well as the model classifying on individual measures of full body movement and general facial expressiveness. This highlights the importance of the social context to capture the manifestation of autistic symptoms.

A model based on reciprocity of facial action units within the interactions showed the best classification accuracy (79.5%) within our sample. When looking more closely at individual feature importance in the facial region, we found heightened and more varied scores for reciprocal adaptation in the AUs chin raiser, jaw drop and lip corner depressor in both tasks to be indicative for classification into the autistic interaction type. This was especially pronounced for the mealplanning task, suggesting higher and more varied synchrony in this task in the ASD-TD interactions. While elevated synchrony in ASD might seem counterintuitive at first glance, especially in light of findings on reduced mimicry in autism ^17,31^, taking a closer look at feature importance for the TD-TD group provides a differentiated picture. Participants with higher values for minimum adaptation across all features had an increased likelihood to be classified into the TD-TD group, suggesting a potential floor effect for facial synchrony in this group. Thus, their synchrony did not subceed a certain lower threshold. This was especially pronounced in the action units for inner brow raiser (AU1), lip stretcher (AU20) and blinking (AU45). Additionally, motor synchrony in autistic interactions has previously been found to vary along with the level of autistic traits, social-communicative functioning, and context ^16^. The same mechanisms may hold true for mimicry. For example, in a study investigating mimicry in the BERT emotion recognition task, Drimalla and colleagues ^32^ found significantly more variance in the intensity of facial expressions in autistic participants. Importantly, since machine learning analyses factor in countless interdependencies between features, interpretations based on feature weights should be considered with caution. Nevertheless, the rather high classification accuracy based solely on facial synchrony features found in our study provides valuable implications for future research on classification based on social interactions in an even more ecological setting (e.g., diagnostic assessments via video conferencing). Interestingly, our model based on measures of individual amount of full body movement and general facial expressiveness (MovEx) was the second-best of the base learners, supporting findings of a characteristic motor signature in autism. For example, Zhao and colleagues ^33^ investigated head movements in autistic children during live interactions and found aberrances on all three axes. Notably though, our classification model factoring in dyad type, thus, data that included the TD interaction partners, showed superior performance compared to classification based on diagnosis. Hence, interactional aspects also seem to have an association with individual movement features, supporting the hypothesis that intra- and interpersonal adjustment processes are not entirely independent of each other ^34^.

Contrary to previous findings of high classification accuracy for head and body coordination ^25^, our model based on this modality performed at a below-chance level, showing low specificity of head-body coordination for autistic vs. non-autistic interaction. However, interpretation should be considered cautiously given the specifications of our experimental setup. Due to our data being collected as part of a larger setup, participants wore wristbands on their non-dominant hand measuring physiological data (see Supplementary Information S1). In order to reduce artefacts in physiological data acquisition, participants were instructed to relax their non-dominant hand in their lap. Arguably, this instruction and setup difference with regards to the previous study could well account for the lack of classification power by intrapersonal coordination in the current study. This is supported by the absence of a significant difference between body synchrony found between our participants’ motion time series and randomly matched time series (see Supplementary Information S2.3).

While our results support previous findings on head motion synchrony as a distinguishing feature of autistic communication ^19^, combining it with facial expression synchrony did not yield a higher prediction accuracy in a stacking model. This was also the case for our overall stacking model. However, stacking may be able to improve predictive performance of any problem primarily in cases where the underlying data is not well represented by a single model ^35^, which is not the case in the current study. Thus, combining several models with significantly different predictive accuracies might in fact harm overall performance of the stacker. Additionally, if the underlying base models are highly correlated, combining them does not necessarily lead to improved performance ^35^. In fact, we did find associations of our MovEx model (total head and body movement and general facial expressiveness) with HEADsync for the ASD-TD group as well with INTRAsync for the TD-TD group (see Supplementary Information S4.5). In our study, we aimed to combine different modalities in a hypothesis-driven way to retain a certain amount of interpretability. We found no added benefit for increasing model complexity. However, it is possible that in order to improve predictive performance of social interactions features, non-verbal aspects of social interaction could be complemented by different modalities in the future, such as speech, eye-movements, physiological or neurological measures. For example, in a recent study conducted by Liao and colleagues ^36^, simultaneous measures of EEG, eye tracking and facial expression were assessed of autistic children viewing social and non-social stimuli. The authors found superior prediction accuracies for the combination of behavioral and physiological classifiers.

Notably, there are several limitations within the scope of the present study.

First, the sample size in the current study is limited. To counter this, we implemented a repeated nested cross-validation structure as well as careful feature reduction methods. Nevertheless, our findings should be considered as proof-of-concept and will have to be validated in a larger sample, possibly including adults with differential diagnoses to examine specificity within a clinical context more closely and, hence, strengthen the translational aspect ^37^. However, we are convinced that the high scalability of our largely automatized setup can facilitate a simplified data collection process within clinical settings.

Second, though interpersonal synchrony has been found to be reduced in interactional dyadic settings independent of partner diagnosis ^20^, a preference for interactions within purely autistic dyads as compared to mixed interactions has been suggested ^11^. This is reflected in theoretical frameworks, such as the “double empathy problem” ^38^ as well as “dialectic misattunement” ^39^, specifying autistic impairments to be especially pronounced between people with fundamentally different ways of information processing and interacting. While this underlines the notion of ASD as a social interaction disorder, in a real-world and especially clinical setting this homogenous combination is rarely to be found, which is why this dyad composition was not assessed in this study.

Third, though highly scalable, we relied on existing computer vision algorithms for our study, which are associated with certain limitations themselves. For example, Motion Energy Analysis (MEA) as a video analysis method has constraints regarding the dimensionality of movement. Because MEA only outputs changes in motion, no specifications regarding direction or magnitude of movement can be made. However, while more distinct investigations of these factors in ASD are certainly desirable, they nevertheless add another layer of complexity to already highly dimensional prediction models and, thus, were not a focus of this study. Regarding facial expression, a range of AUs and participants had to be excluded due to their extent of missing values within their resulting time series. This was partially due to the participants moving out of the camera frame. Though OpenFace employs person-specific normalization by subtracting a “neutral” face from all other frames of a person, the algorithm is nevertheless reported as potentially less accurate if a face does not show a lot of movement dynamics ^40^. However, even considering those technical drawbacks, our FACEsync model achieved high classification accuracy. We believe that with the continuing technological developments within computer vision methodology this limitation will likely be overcome in the future.

Lastly, the application of machine learning in clinical psychology and psychiatry is providing novel possibilities for increased precision in individualized diagnosis, prognosis and treatment ^41^. However, with increasing model complexity, interpretation of findings and their implications become more challenging. While our findings point to the predictive accuracy of reciprocity in social interactions for autism, future research should aim to gain a greater understanding about the underlying mechanisms of those features.

Conclusively, using carefully cross-validated ML algorithms, we were able to classify members of autistic and non-autistic dyads based on multiple objective non-verbal measures of reciprocity in naturalistic social interactions. Facial synchrony within the dyad as unit of analysis ^29^ proved to be the most valuable marker for diagnostic classification of ASD.

## 4. Methods

### 4.1. Sample

We recruited 35 participants with ASD from a clinical database, as well as local autism networks. The diagnosis (F84.0 or F84.5) had to have been given by a qualified clinical psychologist or psychiatrist according to ICD-10 criteria as confirmed by a full diagnostic report. Inclusion criteria were an age between 18-60 years, normal intelligence (IQ > 70, as measured by an IQ score based on a verbal and non-verbal IQ test ^42^) and no current neurological disorder. Additionally, 69 typically developing (TD) participants with no current or history of psychiatric or neurological disorders or psychotropic medication were recruited. Two ASD participants had to be excluded from the final sample because their diagnosis could not be verified on the basis of an incomplete diagnostic report. An additional five ASD participants were excluded during the analysis due to data loss from imprecise facial tracking. Due to the dyadic nature of the study, their interactional partners had to be excluded as well. Another TD-TD dyad was excluded due to technical issues during script loading, leading to a final sample of 88 participants. Groups were matched with respect to age and IQ. A chi-square-test of independence revealed no significant association between group membership and gender, χ^2^(1, *N* = 88) = 2.6, *p* = .11. A description of the final sample can be found in *Table 1*. All participants gave written informed consent before study participation and were compensated monetarily afterwards. The study was approved by the ethics committee of the medical faculty of the LMU Munich and in agreement with the Declaration of Helsinki.

**Table 1.**
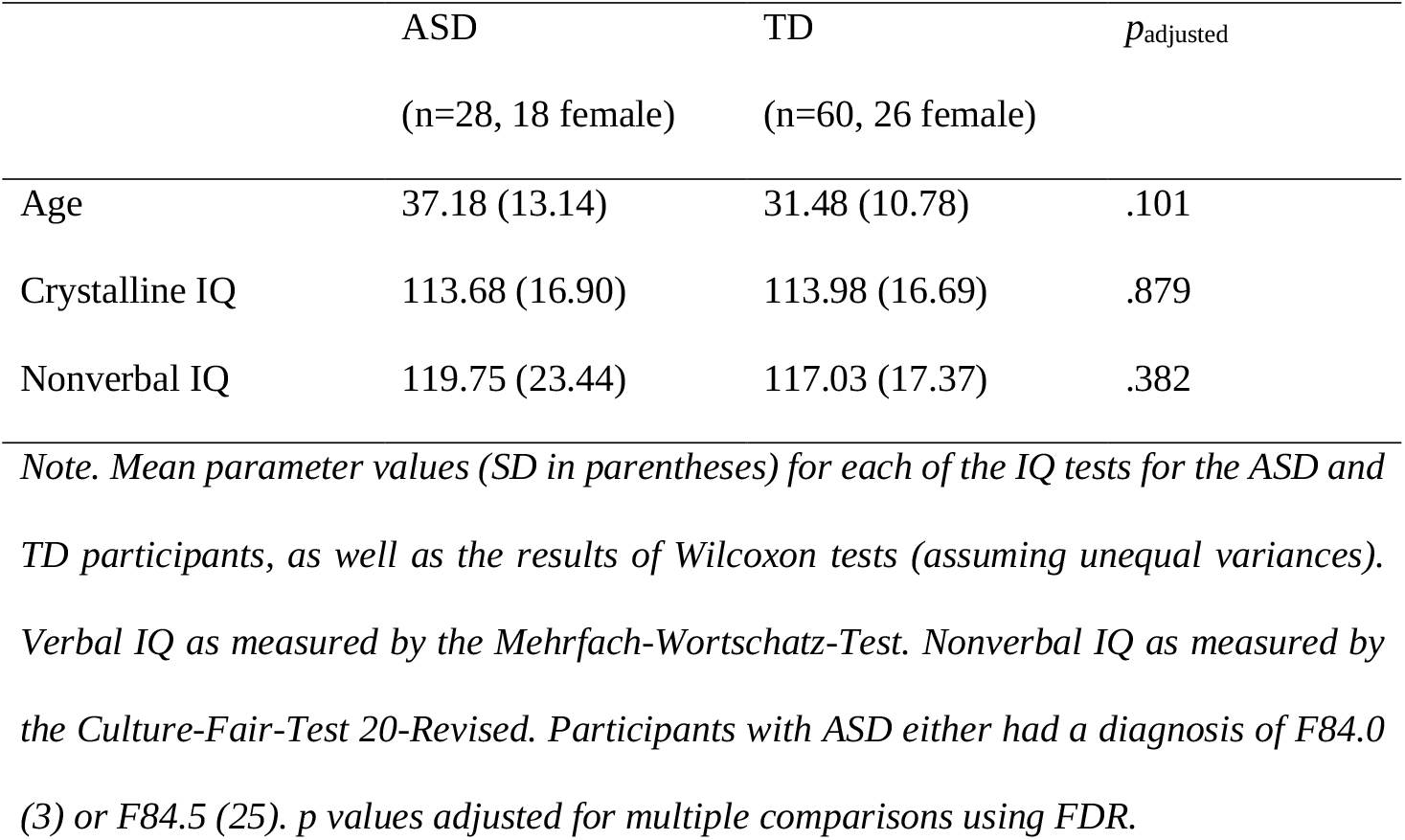
Sample description.

|  | ASD<br>(n=28, 18 female) | TD<br>(n=60, 26 female) | *p*<sub>adjusted</sub> |
| --- | --- | --- | --- |
| Age | 37.18 (13.14) | 31.48 (10.78) | .101 |
| Crystalline IQ | 113.68 (16.90) | 113.98 (16.69) | .879 |
| Nonverbal IQ | 119.75 (23.44) | 117.03 (17.37) | .382 |

*Note. Mean parameter values (SD in parentheses) for each of the IQ tests for the ASD and TD participants, as well as the results of Wilcoxon tests (assuming unequal variances). Verbal IQ as measured by the Mehrfach-Wortschatz-Test. Nonverbal IQ as measured by the Culture-Fair-Test 20-Revised. Participants with ASD either had a diagnosis of F84.0 (3) or F84.5 (25). *p* values adjusted for multiple comparisons using FDR.*

### 4.2. Study setup

Participants were randomly paired resulting in 28 ASD-TD (mixed) and 16 TD-TD (non-autistic control) dyads. All were naïve to the diagnosis of their interactional partner. They were seated approximately 190 cm across from each other in fixed chairs. Two cameras (Logitech C922) were installed on a tripod on a table in front of the participants, recording their respective facial expression at 30 frames per second. A third camera was mounted at a wide angle on a tripod at a distance of approximately 240 cm (Figure 5). All recordings were operated from a single computer using custom PsychoPy ^43^ scripts, allowing for maximal synchronization of the three video input streams. To control for any biases in subsequent video analyses caused by lighting change^44^, measurements were taken in stable artificial light. To maximize hygienic safety measures during the Covid-19 pandemic, slight changes to the setup were required after the first nine participants were assessed (see Supplementary Information S3.3). Participants engaged in two ten-minute conversation tasks for which they were instructed beforehand by the study personnel. After giving a starting cue with a clapping board, all study personnel left the room. Participants were asked to engage in a conversation about their hobbies, as well as to plan a fictional five course meal with dishes they both disliked. The mealplanning task has been used in previous synchrony studies (e.g., ^20,45^), with the rationale that a collaborative task increases affiliation and synchrony respectively. In contrast, a conversation about their hobbies was introduced. Restricted interests are a key diagnostic criterion of autism according to DSM-5 ^1^, whereby autistic individuals tend to switch to monologue style when talking about their interests ^46^ – a unique behavior, which we aimed to capture. The order of the tasks was counterbalanced among participants.

**Figure 5.**
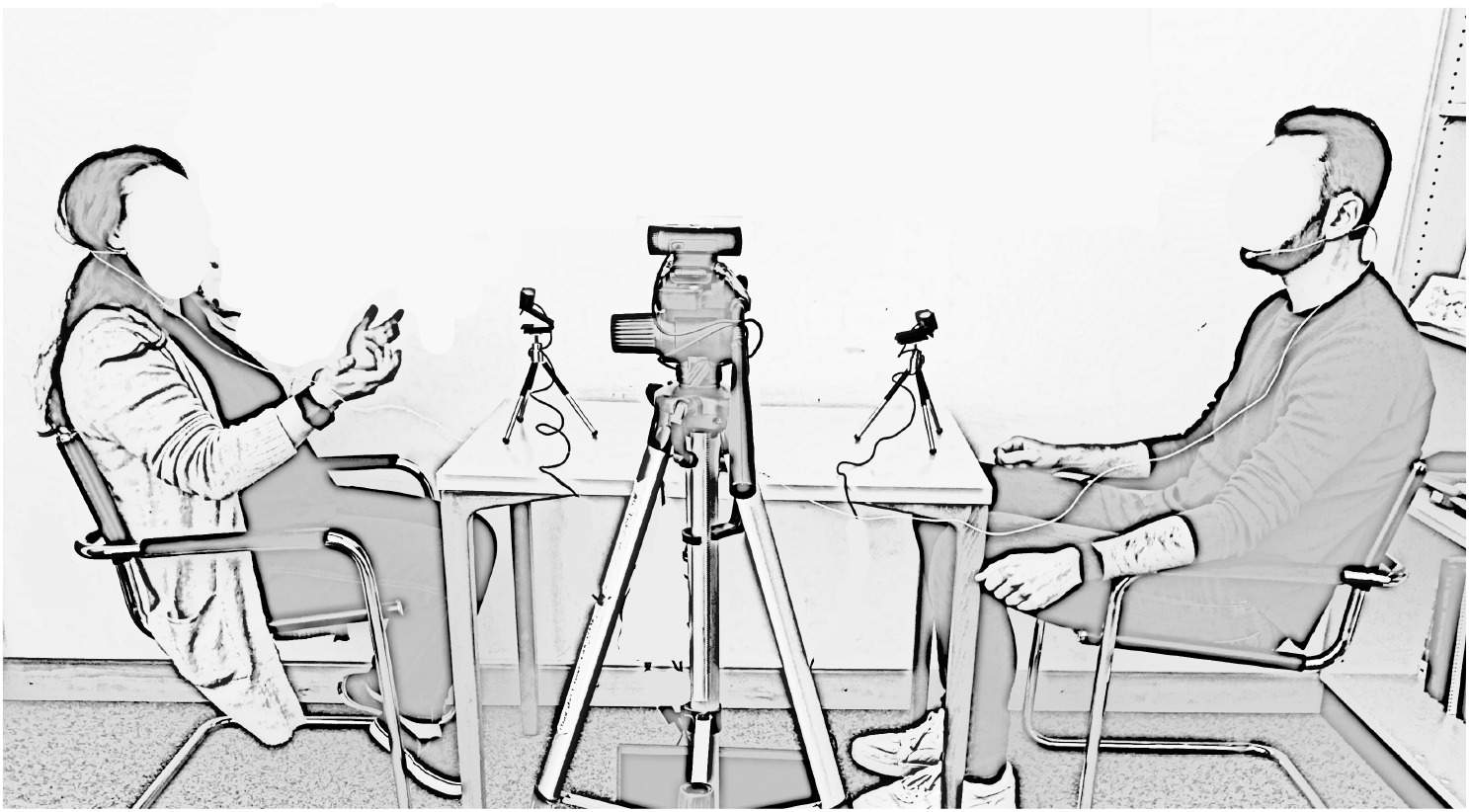
Experimental Setup *Note*. Participants wore head-mounted microphones (t.bone HeadmiKe – D AKG) which were connected to a recorder (Zoom H4N Pro), recording at 44,100 Hz. Additionally, participants were equipped with Empatica E4 29 wristbands on their non-dominant hand. Analyses based on verbal communication and physiological measures are out of the scope of the current study and will be presented elsewhere.

Additionally, participants completed a series of questionnaires to assess their level of self-reported autistic traits (Autism Quotient; AQ^47^), empathy (Saarbrücker Persönlichkeitsfragebogen; SPF ^48^, the German version of the Interpersonal Reactivity Index; IRI ^49^), alexithymia (Toronto Alexithymia Scale; TAS20 ^50^), depressiveness (Beck Depression Inventory; BDI ^51^), self-monitoring (self-monitoring scale; SMS ^52^), and movement difficulties (a German translation of the Adult Dyspraxia Checklist; ADC ^53^). To obtain a best estimate of both their crystalline (Mehrfachwahl-Wortschatz-Intelligenztest; MWT ^54^) and non-verbal (Culture Fair Test; CFT 20-R ^55^) IQ, two IQ assessments were undertaken, and their results averaged. Since difficulties in recognizing emotional facial expressions could potentially cause a bias in the investigation of synchrony in facial expressions, participants additionally completed a computer task for facial expression recognition (Berlin Emotion Recognition Test; BERT ^30^).

### 4.3. Data preparation and feature extraction

Videos were cut to a duration of ten minutes in DaVinci Resolve (Version 16.2.0054). Facial expression was analyzed with the open-source algorithm *Openface 2*.*0* ^56^, identifying action units (AUs) and three head pose parameters (pitch, yaw, roll) and extracting a time series of their presence and intensity for every frame. Motion Energy analysis (MEA ^44^) was used to analyze head and upper body movement captured with the scenic camera. MEA extracts time series of grayscale pixel changes for every frame in pre-specified regions of interest (ROI). Due to the constant lighting conditions and a stable camera, pixel changes within each ROI indicate movement.

Prior to the final analyses, all behavioral time series were synchronized in R using windowed cross-lagged correlation. For this purpose, time series were split into multiple windows, which were subsequently cross-correlated in positive and negative time lags. The size of the respective windows and lags for each modality were carefully chosen, relying on previous research wherever applicable, to ensure maximum standardization. For the estimation of intrapersonal coordination, head movement, as derived from OpenFace, was cross-correlated with the body motion energy times series derived from MEA. Finally, summary scores (mean, median, standard deviation, minimum, maximum, skewness, and kurtosis) of the maximum synchrony instances within both tasks were used as a final feature set. For further details on the cross-correlation and feature extraction procedures, refer to Supplementary Information S1.

Facial emotion recognition capabilities were operationalized as mean accuracy (in %) and response time (in ms) (see Supplementary Information S3.4).

### 4.4. Classification Models

Separate Support Vector Machine (SVM) classification models were trained using features grouped according to the interaction modalities. In each base model, the SVM algorithm independently modeled linear relationships between features and classification label. To account for the interactional nature of the underlying feature set for classification, participants were labelled as belonging to either a mixed (ASD-TD) or non-autistic control (TD-TD) dyad, resulting in groups of 56 and 32 respectively. Linear SVM optimizes a linear hyperplane in a high-dimensional data space that maximizes separability between participants belonging to either of the two dyad types (i.e., the support vectors). Based on the trained hyperplane, the data was subsequently projected into the linear kernel space and their geometric distance to the decision boundary was measured, therefore, predicting each participant’s classification. Every participant was assigned a decision score and a predicted classification label.

We built separate models for the synchrony of facial expression (FACEsync; all qualified AUs in Table 2; 168 features per individual), head movement (HEADsync; global head movement, as well as pitch, yaw and roll; 56 features per individual), and body movement (BODYsync; 14 features per individual), as well as intrapersonal head-body movement coordination (INTRAsync; 14 features per individual), and individual movement parameters (MovEx; total head and body movement, and facial expressiveness; 6 features per individual). The decision scores of all our base models, as well as the model covering the head region (FACEsync + HEADsync), were subsequently combined in a stacking-based data fusion framework ^57^ to assess whether a combination of the modalities would result in superior prediction results than the unimodal classifiers themselves.

**Table 2.** Action Units included in this study as extracted by OpenFace.

| Action Unit | Definition according to FACS |
| --- | --- |
| 1 | Inner brow raiser |
| 2 | Outer brow raiser |
| 6 | Cheek raiser |
| 7 | Lid tightener |
| 9 | Nose wrinkler |
| 14 | Dimpler |
| 15 | Lip corner depressor |
| 17 | Chin raiser |
| 20 | Lip stretcher |
| 23 | Lip tightener |
| 25 | Lips part |
| 26 | Jaw drop |
| 45 | Blink |

*Note. FACS = Facial Action Coding System* <sup>61</sup>

We additionally conducted supplementary analyses using individual diagnosis as classification label. Results of these analyses can be found in the Supplementary Information S3.6.

### 4.5. Support Vector Machine Learning Analysis

Machine learning analyses were conducted with the toolbox NeuroMiner (Version 1.1; https://github.com/neurominer-git/NeuroMiner_1.1) ^58^ in MATLAB (Version 2022b) ^59^. A repeated, nested stratified cross-validation (CV) structure was implemented with 11 outer CV2 folds and ten permutations and ten inner CV1 folds and one permutation. At the outer level, we iteratively held back participants from four dyads as validation samples, while the rest of the data entered the inner cycle, where the data were again split into validation and training sets. Both interactants from a dyad would always remain in the same fold. This nested CV allows for a strict separation between training and testing data, with hyper-parameter tuning happening entirely within the inner loop while the outer loop exclusively measures a model’s generalizability to unseen data. Additionally, the stratified design ensured that proportion of dyad type in every fold would adequately reflect the proportion of dyad type in the full sample. The five base models were preprocessed and trained separately using LIBLINEAR Support Vector L2-regularized L2-loss classification algorithms with linear kernel (see Supplement ary Information S2.1 and S2.2). All models were corrected for class imbalance by hyperplane weighting. Balanced Accuracy (BAC = (sensitivity + specificity)/2) was used as the performance criterion for parameter optimization. Statistical significance of the base classifiers was assessed through permutation testing ^60^, with *α* = 0.05 and 1000 permutations (for details see Supplementary Information S2.3). The two stacking models were trained on the resulting decision scores (all base models, facial expression + head motion synchrony). A L1-loss LIBSVM algorithm with Gaussian kernel was employed to find a parsimonious combination of decision scores which maximized BAC across the C parameter range. For details, see Supplementary Information S3.

## Data Availability

The dataset generated and analyzed during the current study contains clinical information and is therefore not publicly available. They are available from the first author upon reasonable request pending the approval of the coauthors. The preprocessing scripts used during this study are available under https://github.com/jckoe/MLASS-study.

## Acknowledgements

We would like to extend our gratitude to all participants as well as individuals from the autistic community who gave feedback on study design and interpretation during data collection and an outreach event. We would also like to acknowledge Mariia Seleznova for providing valuable advice on ML strategies.

## Author contributions

CFW conceptualized the research program. JCK conceived and programmed the study design. JCK, AN and SF collected the data. JCK and JS pre-processed the data. JCK, MD, ISP, CFW, and NK conceptualized the analysis. MD provided the cross-validation pipeline scripts. JCK analyzed the data and wrote the manuscript. MD, ISP, CFW, and NK provided feedback and consulted on day-to-day issues with JCK. CFW and NK provided the resources. All authors have approved the submitted version.

## Figure legends

*Figure 1. ADC = Adult Dyspaxia Checklist, AQ = Autism Quotient, BDI = Beck Depression Inventory, CFT = Culture Fair Test 20-R, MWT = Mehrfach-Wortschatz-Test, SMS = Self -Monitoring Scale, SPF = Saarbrücker Persönlichkeitsfragebogen, TAS20 = Toronto Alexithymia Scale. The ADC is composed of a section covering movement difficulties in childhood and adulthood. Both scores are depicted in this figure. Bars represent inter-quartile ranges*.

*Figure 5. BAC = balanced accuracy, AUC = area under the curve, PPV = positive predictive value, NPV = negative predictive value. Models are depicted in the order of lowest to highest performing BAC*.

## References

1. American Psychiatric Association. Diagnostic and statistical manual of mental disorders (DSM-5®). (American Psychiatric Pub, 2013).

2. Matson, J. L. & Kozlowski, A. M. The increasing prevalence of autism spectrum disorders. Res Autism Spectr Disord 5, 418–425 (2011).

3. Zwaigenbaum, L. & Penner, M. Autism spectrum disorder: Advances in diagnosis and evaluation. BMJ (Online) vol. 361 Preprint at https://doi.org/10.1136/bmj.k1674 (2018).

4. AWMF. Autismus-Spektrum-Störungen im Kindes-, Jugend-und Erwachsenenalter, Teil 1: Diagnostik: Interdisziplinäre S3-Leitlinie der DGKJP und der DGPPN sowie der beteiligten Fachgesellschaften, Berufsverbände und Patientenorganisationen. https://www.awmf.org/uploads/tx_szleitlinien/028-018l_S3_Autismus-Spektrum-Stoerungen_ASS-Diagnostik_2016-05.pdf (2016).

5. Tariq, Q. et al. Mobile detection of autism through machine learning on home video: A development and prospective validation study. PLoS Med 15, 1–20 (2018).

6. Drimalla, H. et al. Towards the automatic detection of social biomarkers in autism spectrum disorder: introducing the simulated interaction task (SIT). NPJ Digit Med 3, 1–10 (2020).

7. Robles, M. et al. A Virtual Reality Based System for the Screening and Classification of Autism. IEEE Trans Vis Comput Graph 28, 2168–2178 (2022).

8. Redcay, E. et al. Atypical brain activation patterns during a face-to-face joint attention game in adults with autism spectrum disorder. 34, 2511–2523 (2013).

9. Küpper, C. et al. Identifying predictive features of autism spectrum disorders in a clinical sample of adolescents and adults using machine learning. Sci Rep 10, 1–11 (2020).

10. Kosmicki, J. A., Sochat, V., Duda, M. & Wall, D. P. Searching for a minimal set of behaviors for autism detection through feature selection-based machine learning. Transl Psychiatry 5, (2015).

11. Morrison, K. E. et al. Outcomes of real-world social interaction for autistic adults paired with autistic compared to typically developing partners. Autism 1362361319892701 (2019) doi:10.1177/1362361319892701.

12. Sasson, N. J. et al. Neurotypical Peers are Less Willing to Interact with Those with Autism based on Thin Slice Judgments. Sci Rep 7, 40700 (2017).

13. Koehne, S., Hatri, A., Cacioppo, J. T. & Dziobek, I. Perceived interpersonal synchrony increases empathy: Insights from autism spectrum disorder. Cognition 146, 8–15 (2016).

14. Hove, M. J. & Risen, J. L. It’s all in the timing: Interpersonal synchrony increases affiliation. Soc Cogn 27, 949–960 (2009).

15. Miles, L. K., Nind, L. K. & Macrae, C. N. The rhythm of rapport: Interpersonal synchrony and social perception. J Exp Soc Psychol 45, 585–589 (2009).

16. McNaughton, K. A. & Redcay, E. Interpersonal Synchrony in Autism. Curr Psychiatry Rep 22, 12 (2020).

17. Yoshimura, S., Sato, W., Uono, S. & Toichi, M. Impaired Overt Facial Mimicry in Response to Dynamic Facial Expressions in High-Functioning Autism Spectrum Disorders. J Autism Dev Disord 45, 1318–1328 (2015).

18. Zampella, C. J., Bennetto, L. & Herrington, J. D. Computer Vision Analysis of Reduced Interpersonal Affect Coordination in Youth With Autism Spectrum Disorder. Autism Research 13, 2133–2142 (2020).

19. Koehler, J. C. et al. Brief Report: Specificity of Interpersonal Synchrony Deficits to Autism Spectrum Disorder and Its Potential for Digitally Assisted Diagnostics. J Autism Dev Disord (2021) doi:10.1007/s10803-021-05194-3.

20. Georgescu, A. L. et al. Reduced nonverbal interpersonal synchrony in autism spectrum disorder independent of partner diagnosis: a motion energy study. Mol Autism 11, 1–14 (2020).

21. McAuliffe, D., Pillai, A. S., Tiedemann, A., Mostofsky, S. H. & Ewen, J. B. Dyspraxia in ASD: Impaired coordination of movement elements. Autism Res 10, 648–652 (2017).

22. Jansiewicz, E. M. et al. Motor signs distinguish children with high functioning autism and Asperger’s syndrome from controls. J Autism Dev Disord 36, 613–621 (2006).

23. de Marchena, A. & Eigsti, I. M. Conversational gestures in autism spectrum disorders: asynchrony but not decreased frequency. Autism Res 3, 311–322 (2010).

24. Melo, C. et al. Motor stereotypies in autism spectrum disorder: Clinical randomized study and classification proposal. Autism (2022) doi:10.1177/13623613221105479.

25. Georgescu, A. L. et al. Machine Learning to Study Social Interaction Difficulties in ASD. Front Robot AI 6, 1–7 (2019).

26. Stagg, S. D., Slavny, R., Hand, C., Cardoso, A. & Smith, P. Does facial expressivity count? How typically developing children respond initially to children with autism. Autism 18, 704–711 (2014).

27. Cook, J. L., Blakemore, S. J. & Press, C. Atypical basic movement kinematics in autism spectrum conditions. Brain 136, 2816–2824 (2013).

28. Wang, L. A. L., Petrulla, V., Zampella, C. J., Waller, R. & Schultz, R. T. Gross Motor Impairment and Its Relation to Social Skills in Autism Spectrum Disorder: A Systematic Review and Two Meta-Analyses. Psychol Bull 148, 273–300 (2022).

29. Redcay, E. & Schilbach, L. Using second-person neuroscience to elucidate the mechanisms of social interaction. Nat Rev Neurosci 20, 495–505 (2019).

30. Drimalla, H. & Dziobek, I. Berlin Emotion Recognition Test (BERT). (2019).

31. McIntosh, D. N., Reichmann-Decker, A., Winkielman, P. & Wilbarger, J. L. When the social mirror breaks: deficits in automatic, but not voluntary, mimicry of emotional facial expressions in autism. Dev Sci 9, 295–302 (2006).

32. Drimalla, H., Baskow, I., Behnia, B., Roepke, S. & Dziobek, I. Imitation and recognition of facial emotions in autism: a computer vision approach. Mol Autism 12, (2021).

33. Zhao, Z. et al. Atypical Head Movement during Face-to-Face Interaction in Children with Autism Spectrum Disorder. Autism Research 1–12 (2021) doi:10.1002/aur.2478.

34. Bloch, C., Vogeley, K., Georgescu, A. L. & Falter-Wagner, C. M. INTRApersonal Synchrony as Constituent of INTERpersonal Synchrony and Its Relevance for Autism Spectrum Disorder. Front Robot AI 6, 1–8 (2019).

35. Sagi, O. & Rokach, L. Ensemble learning: A survey. Wiley Interdisciplinary Reviews: Data Mining and Knowledge Discovery vol. 8 Preprint at https://doi.org/10.1002/widm.1249 (2018).

36. Liao, M., Duan, H. & Wang, G. Application of Machine Learning Techniques to Detect the Children with Autism Spectrum Disorder. J Healthc Eng 2022, (2022).

37. Dwyer, D. & Krishnadas, R. Five points to consider when reading a translational machine-learning paper. British Journal of Psychiatry vol. 220 169–171 Preprint at https://doi.org/10.1192/bjp.2022.29 (2022).

38. Milton, D. E. M. On the ontological status of autism: the ‘double empathy problem’. Disabil Soc 27, 883–887 (2012).

39. Bolis, D., Balsters, J., Wenderoth, N., Becchio, C. & Schilbach, L. Beyond Autism: Introducing the Dialectical Misattunement Hypothesis and a Bayesian Account of Intersubjectivity. Psychopathology 50, 355–372 (2018).

40. Baltrušaitis, T., Robinson, P. & Morency, L.-P. OpenFace: an open source facial behavior analysis toolkit. https://www.omron.com/ecb/products/mobile/ (2016).

41. Dwyer, D. B., Falkai, P. & Koutsouleris, N. Machine Learning Approaches for Clinical Psychology and Psychiatry. Annu Rev Clin Psychol 14, 91–118 (2018).

42. Schuwerk, T., Vuori, M. & Sodian, B. Implicit and explicit Theory of Mind reasoning in autism spectrum disorders: The impact of experience. Autism 19, 459–468 (2015).

43. Peirce, J. et al. PsychoPy2: Experiments in behavior made easy. Behav Res Methods 51, 195–203 (2019).

44. Ramseyer, F. T. Motion energy analysis (MEA): A primer on the assessment of motion from video. J Couns Psychol 67, 536–549 (2020).

45. Tschacher, W., Rees, G. M. & Ramseyer, F. Nonverbal synchrony and affect in dyadic interactions. 5, (2014).

46. Nadig, A., Lee, I., Singh, L., Bosshart, K. & Ozonoff, S. How does the topic of conversation affect verbal exchange and eye gaze? A comparison between typical development and high-functioning autism. Neuropsychologia 48, 2730–2739 (2010).

47. Baron-Cohen, S., Wheelwright, S., Skinner, R., Martin, J. & Clubley, E. The autism-spectrum quotient (AQ): Evidence from asperger syndrome/high-functioning autism, malesand females, scientists and mathematicians. J Autism Dev Disord 31, 5–17 (2001).

48. Paulus, C. Der Saarbrücker Persönlichkeitsfragebogen SPF (IRI) zur Messung von Empathie: Psychometrische Evaluation der deutschen Version des Interpersonal Reactivity Index. (2009).

49. Davis, M. H. Interpersonal reactivity index. (1980) doi:https://doi-org.emedien.ub.uni-muenchen.de/10.1037/t01093-000.

50. Bagby, R. M., Parker, J. D. A. & Taylor, G. J. The twenty-item Toronto Alexithymia scale. Item selection and cross-validation of the factor structure. J Psychosom Res 38, 23–32 (1994).

51. Hautzinger, M., Bailer, M., Worall, H. & Keller, F. BECK-DEPRESSIONS-INVENTAR - Beck Depression Inventory (BDI; Beck, A.T., Ward, C.H., Mendelson, M., Mock, J. & Erbaugh, J., 1961) - German version. Preprint at http://en.wikipedia.org/wiki/Beck_Depression_Inventory (1994).

52. Graf, A. Eine deutschsprachige Version der Self-Monitoring-Skala. Zeitschrift fur Arbeits-und Organisationspsychologie 48, 109–121 (2004).

53. Kirby, A., Edwards, L., Sugden, D. & Rosenblum, S. The development and standardization of the Adult Developmental Co-ordination Disorders/Dyspraxia Checklist (ADC). Res Dev Disabil 31, 131–139 (2010).

54. Lehrl, S., Merz, J., Burkhard, G. & Fischer, S. Mehrfachwahl-Wortschatz-Intelligenztest. MWT-B, Erlangen: Straube (1999).

55. Weiß, R. H. CFT 20-R: grundintelligenztest skala 2-revision. (Hogrefe, 2006).

56. Baltrušaitis, T., Zadeh, A., Lim, Y. C. & Morency, L.-P. Openface 2.0: Facial behavior analysis toolkit. in 2018 13th IEEE international conference on automatic face & gesture recognition (FG 2018) 59–66 (IEEE, 2018).

57. Wolpert, D. H. Stacked Generalization. Neural Networks vol. 5 (1992).

58. Koutsouleris, N., Vetter, C. & Wiegand, A. Neurominer. Preprint at (2022).

59. MATLAB . Preprint at (2022).

60. Golland, P. & Fischl, B. Permutation Tests for Classification: Towards Statistical Significance in Image-Based Studies. LNCS vol. 2732 (2003).

61. Ekman, P. & Friesen, W. v. Facial action coding system. Environmental Psychology & Nonverbal Behavior (1978).

