## Supplementary Information for "Machine Learning Classification Of Autism Spectrum Disorder Based On Reciprocity In Naturalistic Social Interactions"

### S1. Additional Setup Info

Participants wore head-mounted microphones (t.bone HeadmiKe – D AKG) which were connected to a recorder (Zoom H4N Pro), recording at 44,100 Hz. Additionally, participants were equipped with Empatica E4 [1] wristbands on their non-dominant hand. Analyses based on verbal communication and physiological measures are out of the scope of the current study and will be presented elsewhere.

### S2. Feature extraction

This section lines out further details on the feature extraction procedures for our different machine learning models based on facial expression (S2.1), head movement (S2.2), body movement (S2.3), head-body coordination (S2.5), and full body movement and facial expressiveness (S2.6). Additionally, the derivation procedure of individual feature vectors from the synchrony analyses is explicated in S2.4.

#### S2.1. Facial Expression Synchrony

In order to prevent biases by imprecise facial tracking, we only included participants with a mean confidence of tracked frames over 75%, as well as a percentage of successfully tracked frames over 90%. We were interested in the synchrony of action units rather than emotional facial expressions. According to the Facial Action Coding System (FACS [2]), emotional facial expressions are combinations of certain action units. The correspondence between emotion and combination of action units is assumed to be bidirectional, for example, happiness is assumed to be associated with the activation of the action units 6 and 12, and the activation of these action units is assumed to always convey happiness. This, however, is not appropriate for this study: (1) It is possible that an action unit, though present, is not detected by the automated algorithm used in this study, e.g., due to partial occlusion of the face. In this case the emotional facial expression synchrony would erroneously be coded as non-existent. Assessing all action units separately would still capture synchrony of all correctly detected action units. (2) Inferring emotional states from facial expressions has shown to be less straightforward than originally assumed [3]. Moreover, autistic people have been found to demonstrate even less coherence in their emotional expressions and their underlying emotional states[4].

For the cross-correlation of all facial AUs, the time series were split into windows of seven seconds (in accordance with [5]), which were lagged by two seconds respectively. To prevent information loss at window boundaries [6], we performed the cross-correlations in steps of 4 seconds, returning a matrix of 17,908 cross-correlation values per dyad per task. In line with previous synchrony studies using MEA (e.g., [7,8]), all cross-correlation values were Fisher's Z-transformed and converted to absolute numbers. In time windows where no movement was present by either interactant, the cross-correlation was labelled as missing. To avoid overfitting, in our final machine learning analysis we only included action units for which none of the participants had more than 50% of missing data. For the mealplanning task, these were action units 1, 2, 6, 7, 9, 14, 15, 17, 20, 25, 26, and 45. For the hobbies task, we included action units 1, 2, 6, 7, 9, 15, 17, 20, 23, 25, 26, and 45.

#### S2.2. Head Movement Synchrony

To account for the dynamic nature of the interaction tasks [6], the extracted motion energy time series for head movement were synchronized in windows of 30 seconds and 5 second lags. An overlap of 15 seconds was chosen in order to capture instances of synchrony between windows, resulting in a cross-correlation matrix of 11,438 values per dyad per task.

As is common practice in the application of MEA, we assessed whether the synchrony scores derived from the extracted motion energy values were above chance. To this end, we shuffled our datasets to create 500 pseudo-dyads, thus, pairing the time series of two people who had never actually interacted with each other. Using windowed cross-lagged correlation, we subsequently calculated their interpersonal synchrony in the same manner as the real dyads with window sizes of 30 seconds, increments of 15 seconds and lags of 5 seconds. The resulting cross-correlations were averaged across all windows and lags, resulting in one global synchrony value per pseudo-dyad and compared to the averages derived from the real dyads using independent Welch t-tests. Pseudo-synchrony in the head ROI ($M$ = .075, $SD$ = .013) was significantly lower that real head synchrony ($M$ = .081, $SD$ = .018), suggesting the head synchrony found between our participants to be above chance.

#### S2.3. Body Movement Synchrony

Body motion energy time series were processed and synchronized in the same manner as head motion energy. For the comparison to pseudosynchrony, we found a group difference suggesting above chance body synchrony in our interactional dyads (pseudo: $M$ = .087, $SD$ = .016 vs. real: $M$ = .089, $SD$ = .018). However, contrary to our hypothesis this result was not significant.

#### S2.4. Derivation of individual feature vectors and peak-picking – from shared interpersonal synchrony to individual adaptation

One way to establish interpersonal synchrony is through the adaptation of one person to another. All resulting interpersonal synchrony cross-correlation matrices (facial expression, head motion, and body motion) were split according to the direction of the lag (Supplementary Figure S1). This allowed for quantification of the degree of adaptation of every participant within their dyad[9]. Subsequently, a peak-picking algorithm[10,11] was used to extract the maximum adaptation per time window in every task. Summary statistics (mean, median, standard deviation, minimum, maximum, skewness, and kurtosis) of these peak values per task constituted the final feature set.

*Supplementary Figure S1*. Derivation of individual feature vectors from time lagged windowed cross-correlation.

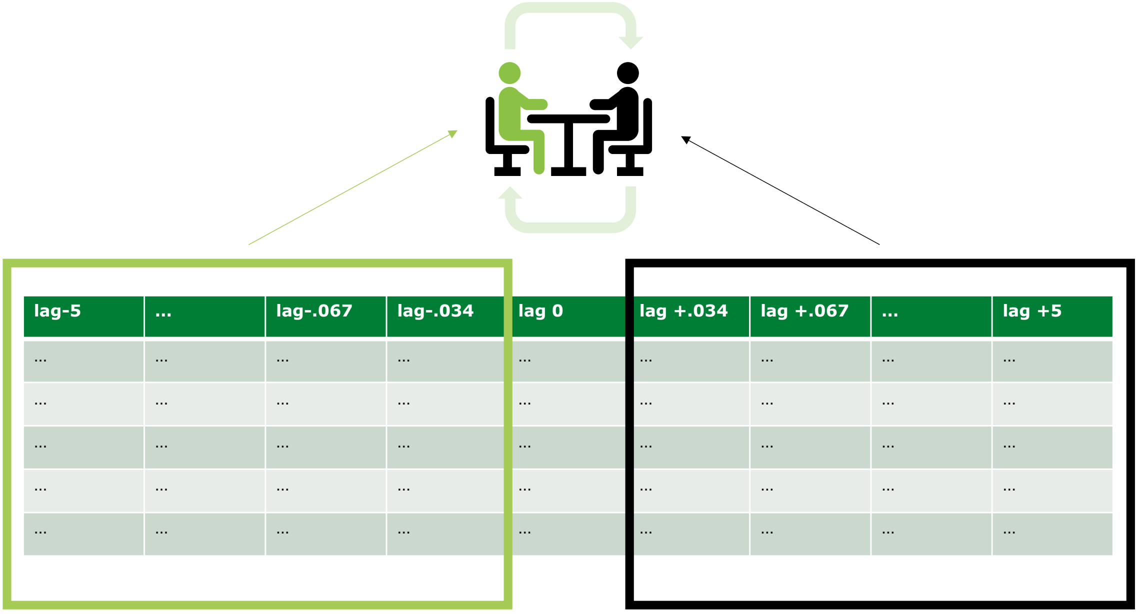


#### S2.5. Head-body coordination

For the purpose of quantifying head and body integration, a three-dimensional head motion vector was derived from the three movement axes (pitch, yaw and roll) using the following formula:

$$head\ movement = \sqrt{{\Delta_{Tx}}^2 + {\Delta_{Ty}}^2 + {\Delta_{Tz}}^2}$$

$\Delta_{Tx}$: frame-to-frame difference vector in head movement on the x-axis with respect to the camera

$\Delta_{Ty}$: frame-to-frame difference vector in head movement on the y-axis with respect to the camera

$\Delta_{Tz}$: frame-to-frame difference vector in head movement towards to and away from the camera

The resulting head movement vector was subsequently cross correlated with the body motion time series derived from MEA. A window size of 30 seconds with lags of 5 seconds and a step size of 15 seconds was chosen in conformity with the calculation of interpersonal movement synchrony. Peak synchrony instances of every time window were extracted, with their summary statistics (mean, median, minimum, maximum, skewness, and kurtosis) per task constituting the final feature set for intrapersonal movement coordination.

#### S2.6. Total Movement and Facial Expressiveness

We aimed to quantify both relative amount of head and body movement and overall facial expressiveness of the individual participants during our testing sessions. Movement quantity of head and body ROI was derived from the respective MEA time series of the two tasks. Following procedures from previous MEA publications [12,13], movement quantity was defined as the number of frames with changes in motion energy divided by the total number of frames, resulting in four values per participant. Facial expressiveness was operationalized as mean intensity of all action units derived from OpenFace 2.0[14] included in our facial expression classifier (AU time series with <50% missing values) per task, resulting in two features per participant.

### S3. Machine learning specifications

#### S3.1. Machine Learning Preprocessing Pipeline for base models

In a first step, all base models underwent pruning of uninformative features (features with 0 variance). Subsequently, all features were scaled from 0 to 1 to remove potential effects of scale differences. Due to the relatively large amounts of features, further pre-processing was conducted on the base models of facial expression synchrony and head movement synchrony.

The base models of facial expression synchrony and head movement synchrony underwent the following additional processing:

(1) To reduce the dimensionality, features were pre-processed using principal component analysis (PCA), retaining the principal components that explained 80% of the variance in each CV1 fold [15].

(2) Subsequently, features were scaled again between 0 and 1.

For all models, the slack parameter was optimized in the inner CV cycle using 11 parameters within the following range: 0.0156, 0.0312, 0.0625, 0.1250, 0.2500, 0.5000, 1, 2, 4, 8, and 16. These represent the default parameter settings in Neurominer [15]. An ensemble of the top 50% performing models was created for each base learner that was subsequently applied to the outer CV2 data to produce a single average robust prediction.

#### S3.2. Machine Learning Preprocessing Pipeline for Stacking Models

We trained two different stacking models to investigate if a combination of modalities in the head region, as well as a combination of all base classifiers could improve prediction accuracy even further. The stacking models combined decision scores of the respective base classifiers (FACEsync + HEADsync; FACEsync + HEADsync + BODYsync + INTRAsync + MoveEx) within each CV1 partition, standardizing the resulting matrices and subsequently using them as new sets of predictive features, which replaced the original features in each CV1 partition. Subsequently, the CV2 validation predictions of the previously trained base classifiers' SVM ensembles were combined and standardized using the median and winsorized within 3 standard deviations to their closest percentile. Then, each SVM ensemble was applied to this standardized CV2 decision score matrix. Majority voting was used to achieve class prediction.

#### S3.3. Permutation testing description

We employed permutation testing to assess whether our base prediction models were statistically significant[16]. To this end, we performed 1000 random permutations of the outcome labels (ASD-TD and TD-TD). All linear SVM models were retrained for each permutation in the same stratified repeated nested double CV using the respective feature subsets obtained from the observed-label analyses. Subsequently, we accumulated the predictions of the random models for each permutation into a permuted ensemble prediction for each outer cycle subject. Thus, we built a null distribution of out-of-training classification performance (BAC) for every base as well as the two stacking classifiers. We then calculated the significance of the observed out-of-training accuracy as the number of events where the permuted out-of-training accuracy was higher or equal to the observed BAC divided by the number of permutations performed. The significance of each model was determined at $\alpha$=0.05, FDR corrected.

#### S3.4. Feature visualization of facial expression features

To determine the influence of the different features on the predictive BAC on the individual level, features were visualized for the best-performing model (FACEsync). Namely, this included the calculation of the weights of the individual features, the cross-validation ratio, and the sign-based consistency. The feature weights (Supplementary Figure S2) calculated by Neurominer were defined as the median weights of the selected CV1 models for each CV2 fold divided by the number of CV2 folds [15]. Cross-validation ratio, a measure of stability, was defined as the sum across CV2 folds of the CV1 median weights divided by their respective CV1 standard error, all of which was subsequently divided by the number of CV2 folds [17]. The sign-based consistency [18] was defined as the number of times that the sign of each feature (positive or negative) was consistent within an ensemble multiplied by the number of times that the feature was non-zero and calculated according to the following procedure [15]: The measure is between 0 to 1, with 1 representing perfect consistency within the ensemble and 0 if the weights are equally positive and negative or when the feature is omitted with a zero weight. A p-value was then calculated by defining a hypothesis test for the importance score with a null hypothesis of 0. A z-score was calculated as the importance divided by the square root of the variance of the importance scores. A standard p-value was then calculated using a normal cumulative distribution function to choose the right-tailed significance. P-values were corrected using the false-discovery rate.

*Supplementary Figure S2.* Feature weights for FACEsync model.

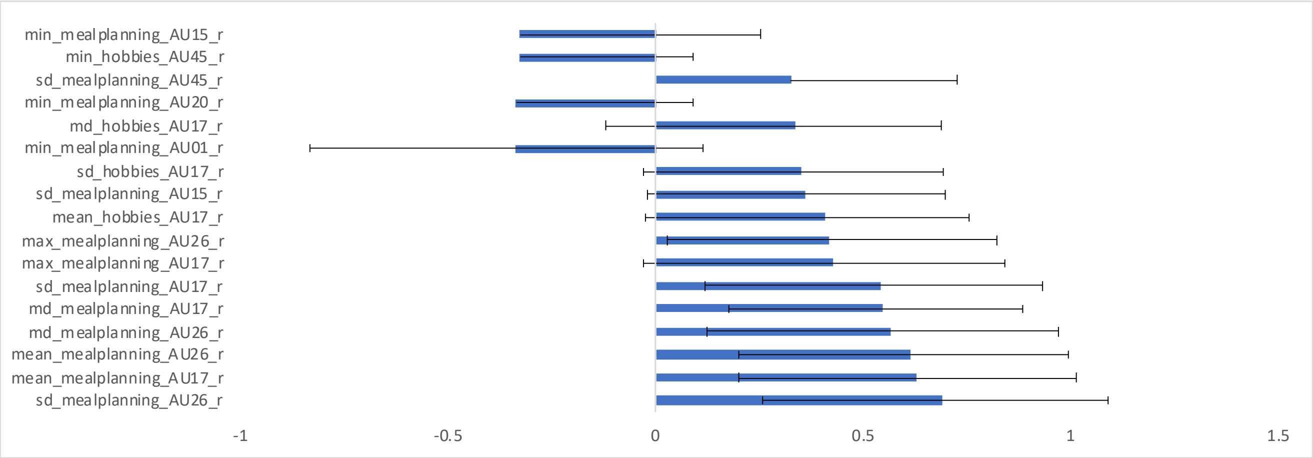


### S4. Supplementary Results

We conducted a range of exploratory analyses to further characterize our sample with standardized clinical self-ratings (S4.1). Moreover, we aimed to explore the impact of our experimental setup on naturalistic interactional behavior of our participants (S4.2 and S4.3). All participants underwent a facial expression recognition classification task whose group-based results can be found under S4.4. To further investigate potential underlying factors driving misclassification within our base models, we conducted additional correlational analyses (S4.5). Lastly, we explored how our interactional setup performed in a series of machine learning classification models based in individual diagnosis (S4.6).

#### S4.1. Clinical self-ratings

We assessed clinical characteristics of our sample using a range of neuro-psychological self-rating questionnaires. Results based on a group comparison between patients and control participants can be found in Supplementary Table S1.

#### S4.2. Camera influence

Since we aimed to capture naturalistic social interactions, we assessed the degree to which the participants felt influenced by the cameras being present. Participants could rate the degree of influence on a 4-point-scale ranging from 0 (= not at all), 1 (= a little), 2 (= considerably), to 3 (= very much). Ratings can be seen found in Supplementary Figure S3. Perceived influence of camera on interaction.

*Supplementary Figure S3.* Perceived influence of camera on interaction.

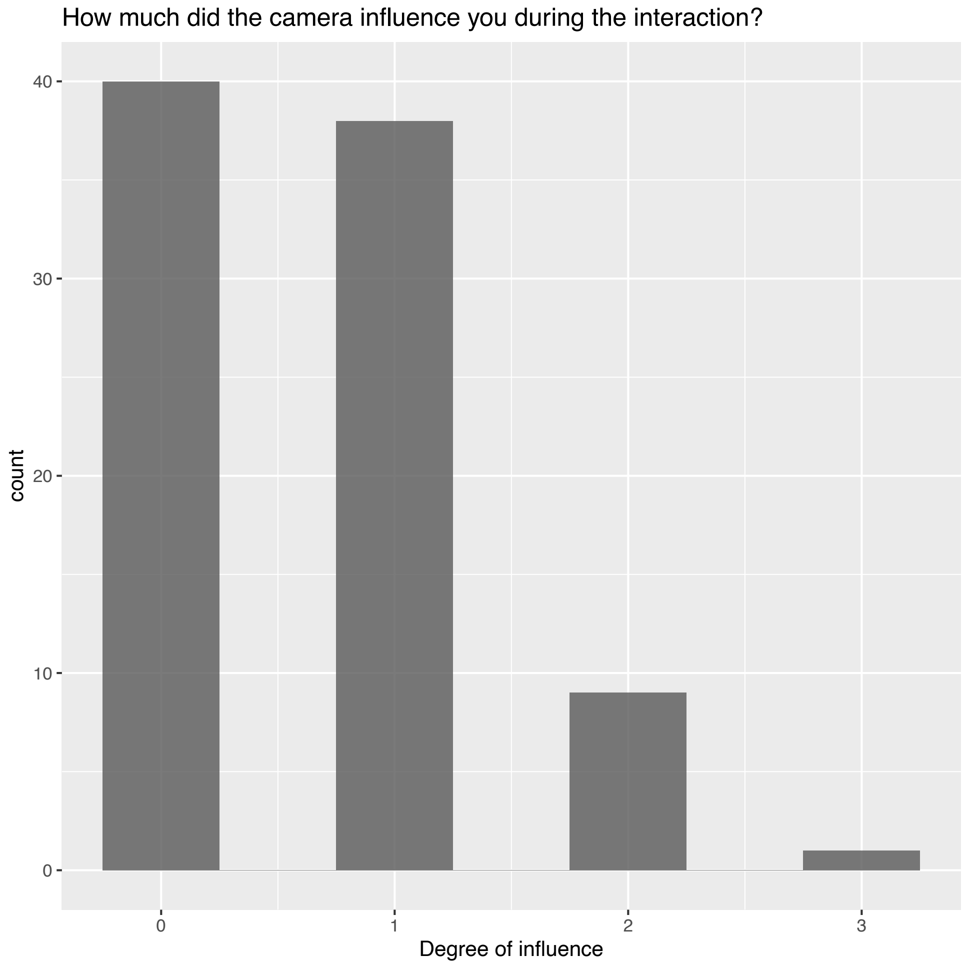


#### S4.3. Impact of COVID-19 measures on interaction

Shortly after the beginning of data collection (the first nine control dyads), the increased hygienic safety measures caused by the Covid-19 pandemic required slight changes in setup. Amongst these changes were a different testing room, as well as the installing of a transparent, plastic screen between the participants to reduce the risk of airborne infections. In an effort to reduce any distracting effects of mirror images on the screen, a transparent anti-reflection foil was applied to it. To rule out any biases regarding those hygienic measures on the features used for classification, we conducted two-sample Welch t-tests between the features of the TD-TD dyads before and after the change in setup (Supplementary Table S2). Additionally, after each testing we asked participants to rate the perceived impairment of the plexiglass, as well as the perceived rapport with their conversational partner (Supplementary Figure S4). Analyzing group differences in perceived rapport between the TD-TD dyads did not result in statistically significant differences before and after the installation of the screen (Supplementary Table S2).

*Supplementary Figure S4.* Perceived influence of plexiglass during interaction for all participants.

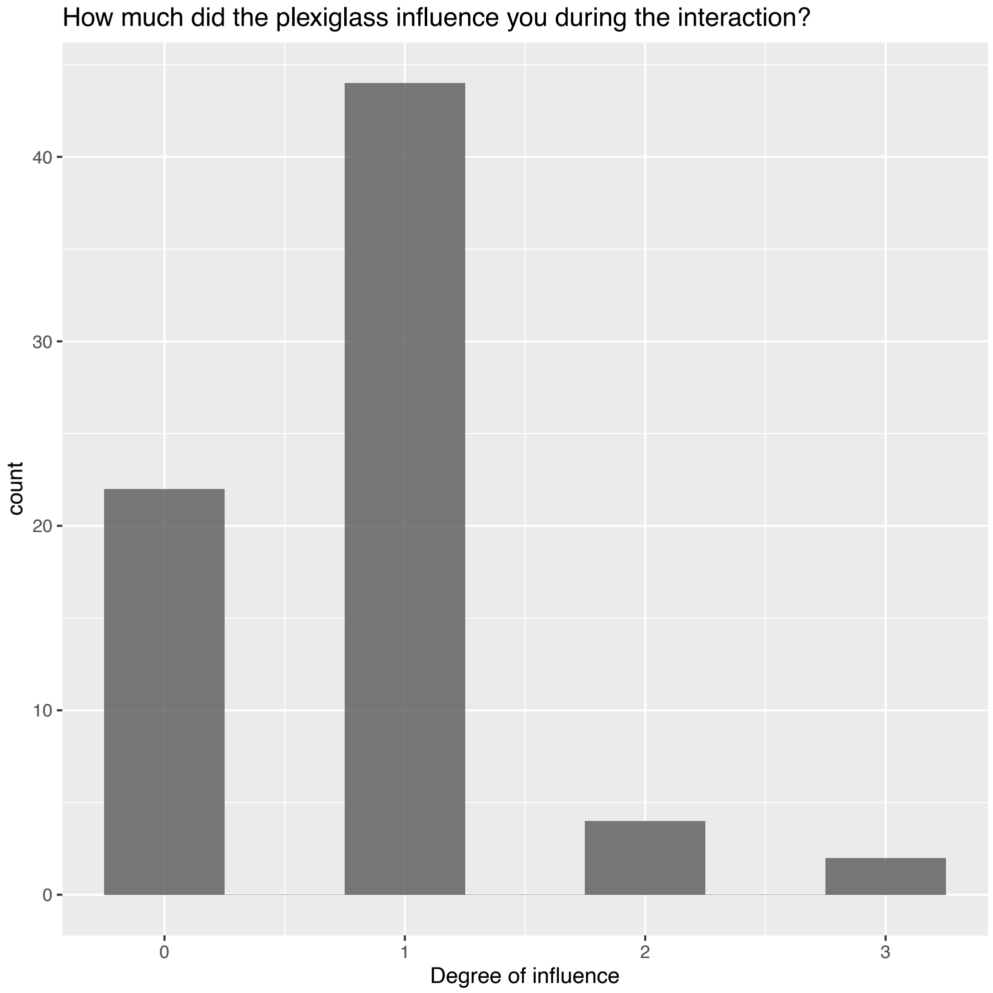


#### S4.4. Facial emotion recognition abilities

Facial emotion recognition abilities were assessed using Berlin Emotion Recognition Test (BERT[19]). In this computer task, participants are shown photos of six different facial expressions of varying intensity in 48 trials (for details see [20]). For every photo, participants decide between one of three alternative emotions that they think is depicted using simple button presses. For each trial, response time and accuracy are recorded. The extracted single scores were averaged across each participant yielding a mean accuracy and response time score per person (see Supplementary Figure S5 for group comparison). In line with previous findings[20], autistic participants responded significantly slower ($M$ = 5141.81 ms, $SD$ = 2380.64 ms) than control participants ($M$ = 3217.31 ms, $SD$ = 1306.17 ms; $t(34.80) = 4.01$, $p_{adj} < .001$). Additionally, autistic participants were able to classify the emotions less accurate ($M$ = .79, $SD$ = .09) than control participants ($M$ = .82, $SD$ = .07). However, this differences did not reach significance ($t(43.34) = -1.64$, $p_{adj} = .11$).

*Supplementary Figure S5.* Comparison of facial expression recognition accuracy and reaction time for diagnostic groups.

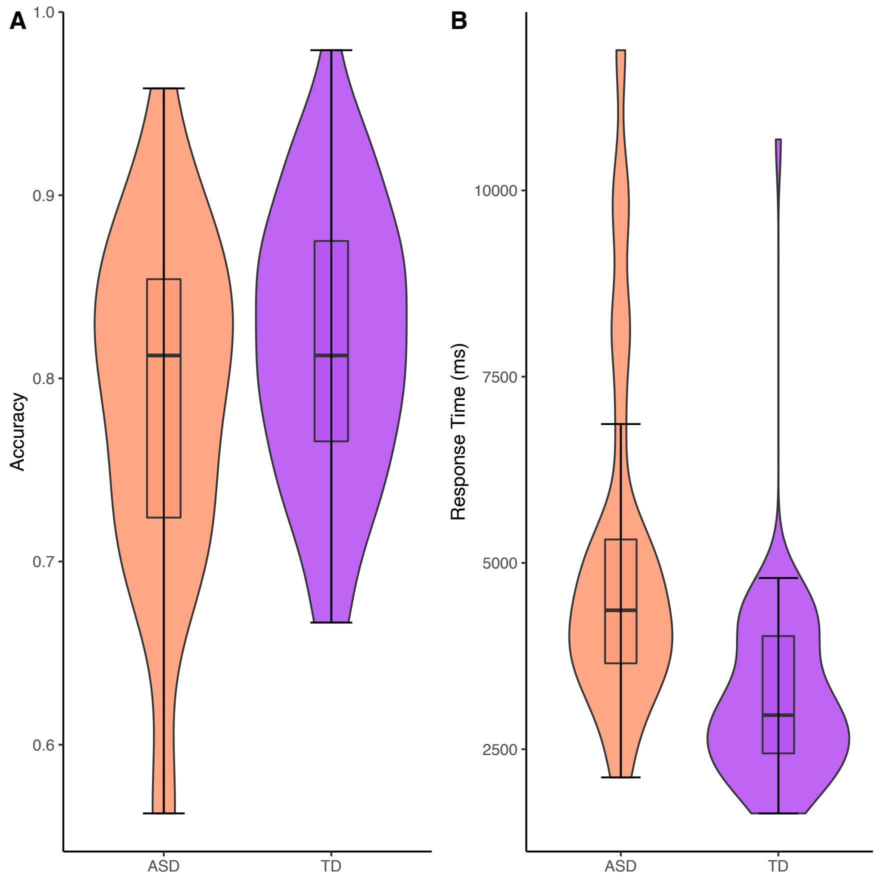

#### S4.5. Correlations between decision scores of base models

To investigate the relationship between the decision scores of the different base models and to examine whether there were associations driving misclassifications in the stacking model, we computed Pearson correlation coefficients within the two groups (ASD-TD and TD-TD). Supplementary Table S3 shows the correlation coefficients for the ASD-TD group, Supplementary Table S4 for the TD-TD group. Supplementary Figure S6 depicts the significant association between the decision scores of our classification model based on total movement and facial expressiveness and the model based on head synchrony for the ASD-TD group. For the TD-TD group, the former was significantly associated with the classification model based on intrapersonal coordination (Supplementary Figure S7).

*Supplementary Figure S6.* Correlation between decision scores of Head Synchrony (HEADsync) and Full Body Movement & Facial Expressiveness (MovEx) Base Models for ASD-TD group

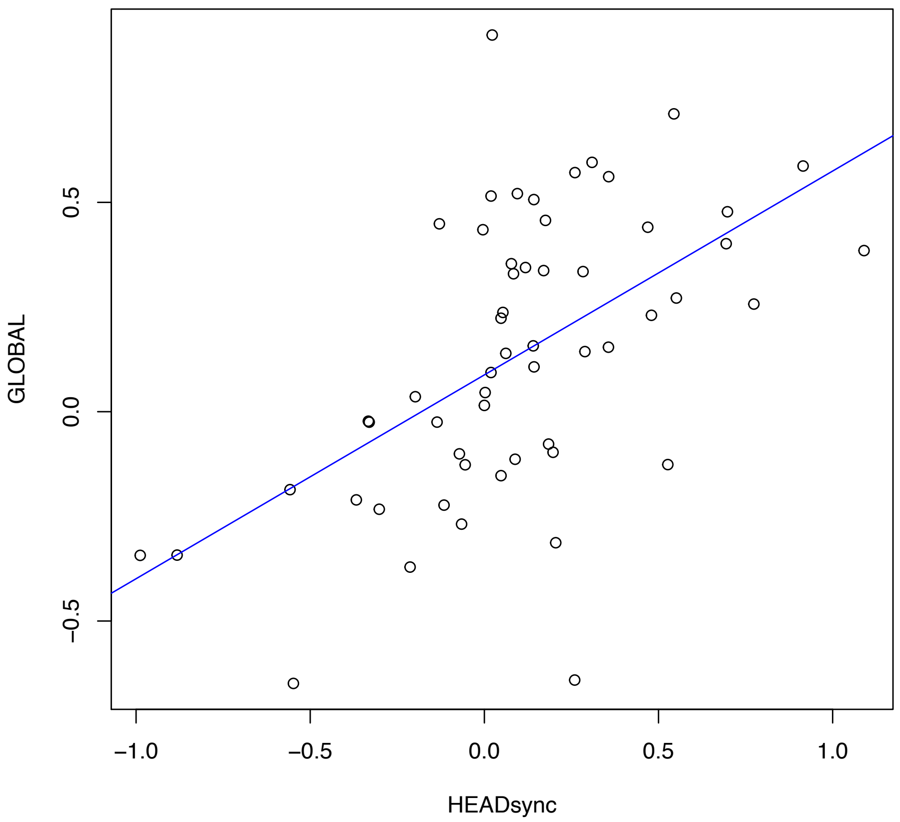

*Supplementary Figure S7.* Correlation between decision scores of intrapersonal head and body coordination (INTRAsync) and Full Body Movement & Facial Expressiveness (MovEx) Base Models for TD-TD group.

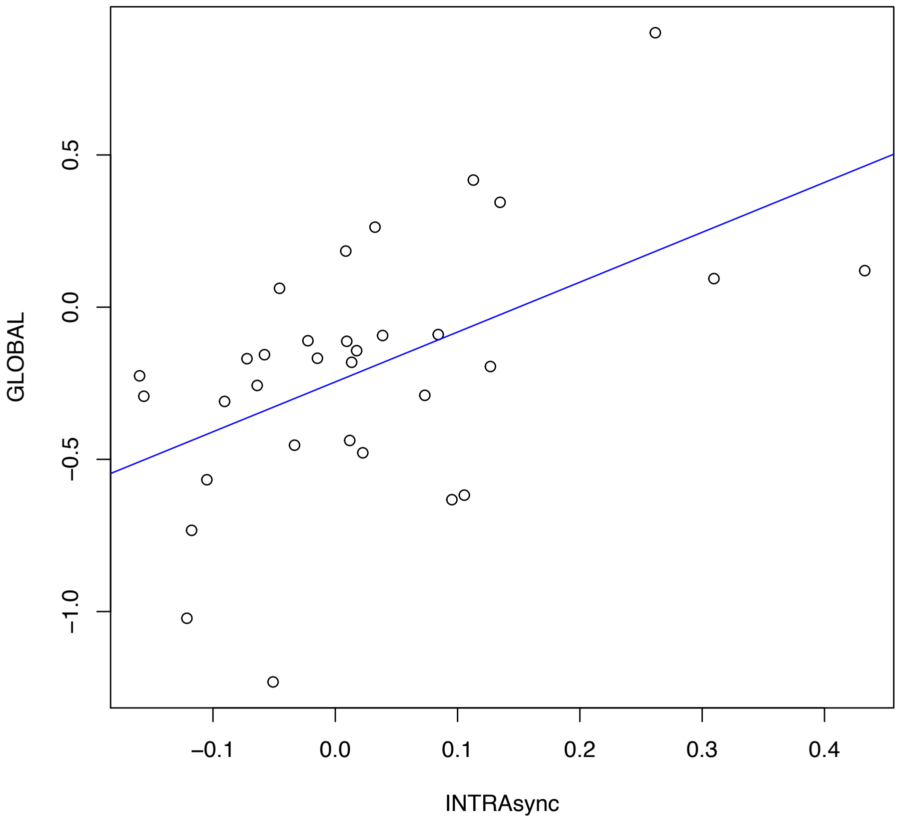


*S4.6. ASD vs. TD analysis results*

To investigate whether our dyadic classification approach outperformed a classification based on diagnostic group while ignoring dyad membership, all SVM analyses were repeated using individual diagnosis as classification label (ASD vs. TD). The repeated, nested stratified CV procedure was adapted accordingly, so that the proportion of autistic and non-autistic participants in every fold reflected the overall proportion within the sample. Once again, both dyad members were always assigned to the same fold in order to prevent information leakage. All models performed around chance level. Detailed classification metrics can be found in Supplementary Table S11.

### S5. Supplementary tables

*Supplementary Table S1. Clinical self-ratings of autistic and control participants.*

| Questionnaire | ASD (n=28, 18 female) | TD (n=60, 26 female) | $p_{adjusted}$ |
|---|---|---|---|
| SPF | 37.32 (7.82) | 45.12 (5.54) | < .001 |
| TAS20 | 59.82 (12.15) | 36.87 (8.88) | < .001 |
| BDI | 14.25 (10.88) | 3.68 (3.65) | < .001 |
| SMS | 6.07 (3.21) | 9.42 (2.98) | < .001 |
| ADC | 47.68 (17.82) | 115.27 (9.30) | < .001 |

*Note. Mean parameter values (SD in parentheses) for each of the questionnaires are shown for the ASD and TD participants, as well as the results of Wilcoxon tests (assuming unequal variance). BDI includes one missing value for TD. AQ = Autism Quotient. SPF = Saarbrücker-Persönlichkeits-Fragebogen. TAS-20 = Toronto Alexithymia Scale. BDI = Beck Depression Inventory. SMS = Self-Monitoring-Scale. ADC = Adult Dyspraxia Checklist. p values adjusted for multiple comparisons using FDR.*

*Supplementary Table S2.* Mean group differences in perceived rapport before and after the Plexiglas setup for TD-TD dyads (n = 16 per group).

| | *M* pre | *M* post | t | *p* | *df* | 95% CI | $p_{adjusted}$ |
|---|---|---|---|---|---|---|---|
| Responsiveness | 2.81 | 2.50 | 1.91 | .067 | 28.33 | [-.02 – .65] | .20 |
| Smoothness | 2.69 | 2.44 | 1.43 | .164 | 29.86 | [-.11 – .61] | .25 |
| Comfort | 2.56 | 2.50 | .34 | .733 | 30.00 | [-.31 – .43] | .73 |

| | *M* pre | *M* post | t | *p* | *df* | 95% CI | $p_{adjusted}$ |
|---|---|---|---|---|---|---|---|

*Note. Two-sample Welch t-test. p-values were adjusted for multiple testing using Bonferroni-Holm. Interaction (= "How responsive did you perceive your interactional partner?"), Smoothness (= "How smooth did you perceive the communication with your partner?"), Comfort (= "How comfortable did you feel during the interaction?"). Ratings went from 0 (= not at all) to 3 (= a lot) on a four-point scale. CI = confidence interval.*

*Supplementary Table S3.* Pearson correlation coefficients between the decision scores of the different base models for the participants from the ASD-TD dyad type

| Model decision scores | *M* | *SD* | 1 | 2 | 3 | 4 |
|---|---|---|---|---|---|---|
| 1. FACEsync | .24 | .37 | | | | |
| 2. HEADsync | .10 | .39 | .09 | | | |
| 3. BODYsync | -.01 | .19 | .01 | .24 | | |
| 4. INTRAsync | -.03 | .17 | .24 | -.01 | .17 | |
| 5. MovEx | .14 | .34 | .15 | .55*** | .01 | .27 |

*Note. M* and *SD* are used to represent mean and standard deviation, respectively. *p* values are FDR corrected. *** indicates $p < .001$.

*Supplementary Table S4.* Pearson correlation coefficients between the decision scores of the different base models for the participants from the TD-TD dyad type

| Model decision scores | *M* | *SD* | 1 | 2 | 3 | 4 |
|---|---|---|---|---|---|---|
| 1. FACEsync | -.30 | .77 | | | | |
| 2. HEADsync | -.01 | .31 | .28 | | | |
| 3. BODYsync | -.02 | .22 | .24 | .37 | | |
| 4. INTRAsync | .02 | .13 | -.14 | .31 | .29 | |
| 5. MoveEx | -.21 | .41 | .25 | .26 | .44 | .52* |

*Note*. *M* and *SD* are used to represent mean and standard deviation, respectively. *p* values are FDR corrected. * indicates $p < .05$.

*Supplementary Table S5. Additional classification metrics for the ASD-TD vs. TD-TD SVM models.*

| Model | True negatives | True positives | False negatives | False positives | Accuracy (%) | Number needed to diagnose | Positive likelihood ratio | Diagnostic odds ratio | Permutation test, *p* value |
|---|---|---|---|---|---|---|---|---|---|
| FACEsync | 24 | 47 | 9 | 8 | 80.7 | 1.7 | 3.4 | 11.3 | < .001 |
| MoveEx | 24 | 35 | 21 | 8 | 67.0 | 2.7 | 2.5 | 6.2 | < .001 |
| HEADsync | 18 | 38 | 18 | 14 | 63.6 | 4.1 | 1.6 | 2.4 | .001 |
| BODYsync | 22 | 25 | 31 | 10 | 53.4 | 7.5 | 1.4 | 2.0 | .011 |
| INTRAsync | 14 | 25 | 31 | 18 | 44.3 | -8.6 | .80 | .6 | .999 |
| FACEsync + HEADsync | 23 | 43 | 13 | 9 | 75.0 | 2.1 | 2.7 | 7.5 | NA |
| All | 23 | 48 | 8 | 9 | 80.7 | 1.7 | 3.0 | 9.3 | NA |

*Supplementary Table S6. FACEsync MODEL: Comparison Analyses of Clinical Variables between Correctly Classified and Misclassified ASD participants.*

| Variables | $M_{correct}$ | $M_{misclassified}$ | $n_{correct}$ | $n_{misclassified}$ | $t$ | $p$ | $df$ | conf.low | conf.high | $p_{adjusted}$ |
|---|---|---|---|---|---|---|---|---|---|---|
| ADC_adult | 34.61 | 35.80 | 23 | 5 | -.15 | .885 | 5.30 | -21.11 | 18.73 | .89 |
| ADC_child | 13.48 | 10.00 | 23 | 5 | 1.16 | .295 | 5.48 | -4.05 | 11.01 | .83 |
| AQ | 34.78 | 34.00 | 23 | 5 | .17 | .874 | 5.27 | -11.14 | 12.71 | .89 |
| BDI | 13.83 | 16.20 | 23 | 5 | -.34 | .745 | 4.88 | -20.26 | 15.51 | .89 |
| CFT_IQ | 121.13 | 113.40 | 23 | 5 | .73 | .488 | 6.62 | -17.48 | 32.95 | .83 |
| MWT_IQ | 114.74 | 108.80 | 23 | 5 | .67 | .530 | 5.59 | -16.16 | 28.03 | .83 |
| SMS_short | 6.35 | 4.80 | 23 | 5 | 1.51 | .155 | 12.98 | -0.66 | 3.76 | .70 |
| SPF_final | 37.87 | 34.80 | 23 | 5 | .64 | .550 | 4.97 | -9.26 | 15.40 | .83 |
| TAS20 | 57.91 | 68.60 | 23 | 5 | -3.06 | .008 | 15.62 | -18.10 | -3.27 | .07 |

*Supplementary Table S7. HEADsync MODEL: Comparison Analyses of Clinical Variables between Correctly Classified and Misclassified ASD participants.*

| Variables | $M_{correct}$ | $M_{misclassified}$ | $n_{correct}$ | $n_{misclassified}$ | $t$ | $p$ | $df$ | conf.low | conf.high | $p_{adjusted}$ |
|---|---|---|---|---|---|---|---|---|---|---|
| ADC_adult | 32.10 | 41.62 | 20 | 8 | -2.09 | .048 | 21.80 | -18.97 | -.08 | .22 |
| ADC_child | 11.25 | 16.88 | 20 | 8 | -2.72 | .016 | 14.46 | -10.04 | -1.21 | .14 |
| AQ | 34.45 | 35.12 | 20 | 8 | -.27 | .790 | 26.00 | -5.84 | 4.49 | .79 |
| BDI | 15.45 | 11.25 | 20 | 8 | .98 | .342 | 14.93 | -4.92 | 13.32 | .69 |
| CFT_IQ | 122.25 | 113.50 | 20 | 8 | .73 | .482 | 9.34 | -18.13 | 35.63 | .69 |
| MWT_IQ | 116.40 | 106.88 | 20 | 8 | 1.38 | .190 | 13.18 | -5.36 | 24.41 | .57 |
| SMS_short | 5.90 | 6.50 | 20 | 8 | -.43 | .672 | 12.56 | -3.60 | 2.40 | .76 |
| SPF_final | 37.80 | 36.12 | 20 | 8 | .63 | .537 | 21.81 | -3.86 | 7.21 | .69 |
| TAS20 | 60.95 | 57.00 | 20 | 8 | .86 | .404 | 16.43 | -5.81 | 13.71 | .69 |

*Supplementary Table S8. BODYsync MODEL: Comparison Analyses of Clinical Variables between Correctly Classified and Misclassified ASD participants.*

| Variables | $M_{correct}$ | $M_{misclassified}$ | $n_{correct}$ | $n_{misclassified}$ | $t$ | $p$ | $df$ | conf.low | conf.high | $p_{adjusted}$ |
|---|---|---|---|---|---|---|---|---|---|---|
| ADC_adult | 35.00 | 34.71 | 11 | 17 | .06 | .957 | 23.57 | -10.75 | 11.34 | .96 |
| ADC_child | 14.91 | 11.53 | 11 | 17 | 1.73 | .096 | 26.00 | -.64 | 7.40 | .86 |
| AQ | 34.00 | 35.06 | 11 | 17 | -.35 | .729 | 25.76 | -7.27 | 5.16 | .94 |
| BDI | 15.36 | 13.53 | 11 | 17 | .44 | .661 | 23.81 | -6.69 | 10.36 | .94 |
| CFT_IQ | 117.18 | 121.41 | 11 | 17 | -.47 | .642 | 23.24 | -22.80 | 14.34 | .94 |
| MWT_IQ | 114.45 | 113.18 | 11 | 17 | .20 | .845 | 23.63 | -12.06 | 14.61 | .95 |
| SMS_short | 5.18 | 6.65 | 11 | 17 | -1.18 | .251 | 20.96 | -4.05 | 1.12 | .94 |
| SPF_final | 38.64 | 36.47 | 11 | 17 | .82 | .420 | 23.92 | -3.28 | 7.61 | .94 |
| TAS20 | 62.45 | 58.12 | 11 | 17 | .94 | .358 | 22.91 | -5.22 | 13.90 | .94 |

*Supplementary Table S9. INTRAsync MODEL: Comparison Analyses of Clinical Variables between Correctly Classified and Misclassified ASD participants.*

| Variables | $M_{correct}$ | $M_{misclassified}$ | $n_{correct}$ | $n_{misclassified}$ | $t$ | $p$ | $df$ | conf.low | conf.high | $p_{adjusted}$ |
|---|---|---|---|---|---|---|---|---|---|---|
| ADC_adult | 29.92 | 39.07 | 13 | 15 | -1.80 | .084 | 25.45 | -19.61 | 1.33 | .29 |
| ADC_child | 13.08 | 12.67 | 13 | 15 | .18 | .855 | 24.33 | -4.18 | 5.00 | .88 |
| AQ | 35.46 | 33.93 | 13 | 15 | .49 | .628 | 25.35 | -4.88 | 7.94 | .88 |
| BDI | 10.69 | 17.33 | 13 | 15 | -1.73 | .097 | 22.16 | -14.59 | 1.31 | .29 |
| CFT_IQ | 119.00 | 120.40 | 13 | 15 | -.15 | .878 | 25.54 | -20.00 | 17.20 | .88 |
| MWT_IQ | 110.85 | 116.13 | 13 | 15 | -.81 | .426 | 23.62 | -18.76 | 8.19 | .88 |
| SMS_short | 6.31 | 5.87 | 13 | 15 | .36 | .721 | 26.00 | -2.07 | 2.96 | .88 |
| SPF_final | 34.15 | 40.07 | 13 | 15 | -2.11 | .045 | 24.70 | -11.70 | -.13 | .29 |
| TAS20 | 61.15 | 58.67 | 13 | 15 | .54 | .596 | 25.96 | -7.03 | 12.00 | .88 |

*Supplementary Table S10. MovEx MODEL: Comparison Analyses of Clinical Variables between Correctly Classified and Misclassified ASD participants.*

| Variables | $M_{correct}$ | $M_{misclassified}$ | $n_{correct}$ | $n_{misclassified}$ | *t* | *p* | *df* | conf.low | conf.high | $p_{adjusted}$ |
|---|---|---|---|---|---|---|---|---|---|---|
| ADC_adult | 34.53 | 35.44 | 19 | 9 | -.17 | .864 | 19.54 | -12.00 | 10.16 | .86 |
| ADC_child | 12.47 | 13.67 | 19 | 9 | -.53 | .602 | 17.59 | -5.92 | 3.54 | .85 |
| AQ | 35.05 | 33.78 | 19 | 9 | .49 | .626 | 25.52 | -4.04 | 6.59 | .85 |
| BDI | 16.37 | 9.78 | 19 | 9 | 1.63 | .119 | 18.54 | -1.87 | 15.05 | .36 |
| CFT_IQ | 118.79 | 121.78 | 19 | 9 | -.31 | .757 | 16.44 | -23.08 | 17.10 | .85 |
| MWT_IQ | 117.05 | 106.56 | 19 | 9 | 1.63 | .121 | 17.27 | -3.05 | 24.04 | .36 |
| SMS_short | 6.26 | 5.67 | 19 | 9 | .45 | .658 | 15.65 | -2.21 | 3.41 | .85 |
| SPF_final | 37.05 | 37.89 | 19 | 9 | -.36 | .723 | 23.13 | -5.66 | 3.99 | .85 |
| TAS20 | 62.95 | 53.22 | 19 | 9 | 2.18 | .043 | 17.44 | .34 | 19.11 | .36 |

*Supplementary Table S11*. Classification metrics based on ASD vs. TD classification

| Model ASD vs. TD | BAC (%) | AUC | Sens. (%) | Spec. (%) | PPV (%) | NPV (%) | TN | TP | FN | FP | Acc. (%) | # needed to diagnose | Positive likelihood ratio | Diagnostic odds ratio | Permutation test, *p* value |
|---|---|---|---|---|---|---|---|---|---|---|---|---|---|---|---|
| FACEsync | 55.5 | .55 | 39.3 | 71.7 | 39.3 | 71.7 | 43 | 11 | 17 | 17 | 61.4 | 9.1 | 1.4 | 1.9 | .041 |
| HEADsync | 53.9 | .59 | 42.9 | 65.0 | 36.4 | 70.9 | 39 | 12 | 16 | 21 | 58.0 | 12.7 | 1.2 | 1.5 | .065 |
| FACEsync + HEADsync | 60.2 | .57 | 57.1 | 63.3 | 42.1 | 76.0 | 38 | 16 | 12 | 22 | 61.4 | 4.9 | 1.6 | 2.4 | NA |
| BODYsync | 49.5 | .49 | 35.7 | 63.3 | 31.2 | 67.9 | 38 | 10 | 18 | 22 | 54.5 | -105.0 | 1.0 | .9 | .639 |
| INTRAsync | 38.3 | .35 | 25.0 | 51.7 | 19.4 | 59.6 | 31 | 7 | 21 | 29 | 43.2 | -4.3 | .5 | .3 | 1 |
| MoveEx | 66.1 | .69 | 57.1 | 75.0 | 51.6 | 78.9 | 45 | 16 | 12 | 15 | 69.3 | 3.1 | 2.3 | 5.2 | < .001 |
| ALL | 63.6 | .64 | 57.1 | 70.0 | 47.1 | 77.8 | 42 | 16 | 12 | 18 | 65.9 | 3.7 | 1.9 | 3.6 | NA |

### Supplementary References


1. Empatica E4. https://www.empatica.com/research/e4/.
2. Ekman, P. & Friesen, W. v. Facial action coding system. *Environmental Psychology & Nonverbal Behavior* (1978).
3. Durán, J. I., Reisenzein, R. & Fernández-Dols, J.-M. Coherence between emotions and facial expressions. *The science of facial expression* 107–129 (2017).
4. Costa, A. P., Steffgen, G. & Samson, A. C. Expressive incoherence and alexithymia in autism spectrum disorder. *J Autism Dev Disord* **47**, 1659–1672 (2017).
5. Zampella, C. J., Bennetto, L. & Herrington, J. D. Computer Vision Analysis of Reduced Interpersonal Affect Coordination in Youth With Autism Spectrum Disorder. *Autism Research* **13**, 2133–2142 (2020).
6. Schoenherr, D. *et al.* Quantification of nonverbal synchrony using linear time series analysis methods: Lack of convergent validity and evidence for facets of synchrony. *Behav Res Methods* **51**, 361–383 (2019).
7. Zadok, E., Gordon, I., Navon, R., Rabin, S. J. & Golan, O. Shifts in Behavioral Synchrony in Response to an Interaction Partner's Distress in Adolescents With and Without ASD. *J Autism Dev Disord* (2021) doi:10.1007/s10803-021-05307-y.
8. Georgescu, A. L. *et al.* Reduced nonverbal interpersonal synchrony in autism spectrum disorder independent of partner diagnosis: a motion energy study. *Mol Autism* **11**, 1–14 (2020).
9. Ramseyer, F. *et al.* Exploring nonverbal synchrony in borderline personality disorder: A double-blind placebo-controlled study using oxytocin. *British Journal of Clinical Psychology* **n/a**, (2019).
10. Schoenherr, D. *et al.* Identification of movement synchrony: Validation of windowed cross-lagged correlation and -regression with peak-picking algorithm. *PLoS One* **14**, 1–24 (2019).
11. Boker, S. M., Xu, M., Rotondo, J. L. & King, K. Windowed cross-correlation and peak picking for the analysis of variability in the association between behavioral time series. *Psychol Methods* **7**, 338–355 (2002).
12. Koehler, J. C. *et al.* Brief Report: Specificity of Interpersonal Synchrony Deficits to Autism Spectrum Disorder and Its Potential for Digitally Assisted Diagnostics. *J Autism Dev Disord* (2021) doi:10.1007/s10803-021-05194-3.
13. Ramseyer, F. & Tschacher, W. Nonverbal synchrony of head- and body-movement in psychotherapy: different signals have different associations with outcome. *Front Psychol* **5**, 979 (2014).
14. Baltrušaitis, T., Zadeh, A., Lim, Y. C. & Morency, L.-P. Openface 2.0: Facial behavior analysis toolkit. in *2018 13th IEEE international conference on automatic face & gesture recognition (FG 2018)* 59–66 (IEEE, 2018).
15. Koutsouleris, N., Vetter, C. & Wiegand, A. Neurominer. Preprint at (2022).
16. Golland, P. & Fischl, B. *Permutation Tests for Classification: Towards Statistical Significance in Image-Based Studies*. *LNCS* vol. 2732 (2003).
17. Koutsouleris, N. *et al.* Prediction Models of Functional Outcomes for Individuals in the Clinical High-Risk State for Psychosis or with Recent-Onset Depression: A Multimodal, Multisite Machine Learning Analysis. *JAMA Psychiatry* **75**, 1156–1172 (2018).
18. Gómez-Verdejo, V., Parrado-Hernández, E. & Tohka, J. Sign-Consistency Based Variable Importance for Machine Learning in Brain Imaging. *Neuroinformatics* **17**, 593–609 (2019).

19. Drimalla, H. & Dziobek, I. Berlin Emotion Recognition Test (BERT). (2019).
20. Drimalla, H., Baskow, I., Behnia, B., Roepke, S. & Dziobek, I. Imitation and recognition of facial emotions in autism: a computer vision approach. *Mol Autism* **12**, 1–15 (2021).
